# Predicting the timing of first sustained cognitive worsening in Alzheimer’s disease using real-world clinical data and machine learning

**DOI:** 10.64898/2026.06.02.26354764

**Authors:** Shruthi Venkatesh, Sinian Zhang, Wen Zhu, Michele Morris, Rocco Mercurio, Sarah B Berman, Hansruedi Mathys, Abby L Olsen, C. Elizabeth Shaaban, Shyam Visweswaran, Oscar L Lopez, Tianxi Cai, Jue Hou, Zongqi Xia

## Abstract

**Background:** Cognitive assessments are sparsely documented in electronic health records (EHRs), limiting scalable detection of cognitive worsening in real-world clinical settings.

**Methods:** We applied a deep neural network optimized for identifying clinical event timing from sparsely labeled gold-standard data (*label-efficient incident phenotyping from longitudinal EHR*, LATTE) to predict time-to-first sustained cognitive worsening in AD patients from a large healthcare system (2011–2022) with linkage to an AD Research Center registry in a subset. Sustained cognitive worsening was defined as cognitive decline persisting over ≥2 consecutive visits within 3 years. Separate LATTE models were trained with worsening labels from Clinical Dementia Rating (CDR), Mini-Mental Status Examination (MMSE), and Montreal Cognitive Assessment (MoCA) scores; semi-supervised learning scaled predictions to larger imputation cohorts lacking sufficient longitudinal scores. We evaluated model performance using average time-specific area under the receiver operating characteristic curve (AUC), area between curves (ABC), and Brier scores. To demonstrate clinical utility, we examined whether predicted time-to-worsening differentiated clinically meaningful patient subgroups using competing-risk Cox proportional hazards models accounting for death.

**Findings:** The cohort comprised 27,614 AD patients (65% women, 91% non-Hispanic White, mean [SD] age at start of follow-up 78.76 [9.53] years). In gold-standard cohorts (n: CDR=632, MMSE=710, MoCA=752; remaining patients formed imputation cohorts), LATTE demonstrated robust predictive performance (average time-AUC: CDR 0.816, MMSE 0.694, MoCA 0.710; ABC: CDR 0.067, MMSE 0.293, MoCA 0.078; Brier score: CDR 0.252, MMSE 0.437, MoCA 0.295). *APOE*-ε4 carriers had shorter predicted time-to-worsening compared to non-carriers across all assessments in the imputation cohorts (HRs 1.241–1.376, all *p*<0.025), and k-means derived patient clusters showed differential time-to-worsening in the overall and imputation cohorts (HRs 0.777–0.908, all *p*<.001).

**Interpretation:** LATTE enables *scalable* prediction of sustained cognitive worsening timing, differentiating clinically meaningful patient subgroups. This approach could improve AD clinical monitoring and decision-making in routine care and support targeted clinical trial enrichment.

**RESEARCH IN CONTEXT:** 

**Evidence before this study:** The growing burden of Alzheimer’s disease (AD) creates an urgent unmet need for pragmatic tools to monitor cognitive decline at the point of care and identify target patient populations for clinical trial recruitment. However, cognitive assessments are sparsely documented in electronic health records (EHRs), and fluctuating scores can obscure true worsening, whereas specialized fluid and neuroimaging biomarkers are rarely available outside research settings, limiting scalable real-world utility.

**Added value of this study:** We applied a deep neural network algorithm optimized for identifying clinical event timing from sparsely labeled longitudinal EHR data (LATTE) to predict time-to-first sustained cognitive worsening across three complementary cognitive assessments in AD patients from a large healthcare system: the Clinical Dementia Rating (CDR), Mini-Mental State Examination (MMSE), and Montreal Cognitive Assessment (MoCA). We defined sustained cognitive worsening as clinically meaningful decline (without improvement) persisting over ≥2 consecutive visits within 3 years. We trained LATTE on gold-standard cohorts with sparse outcome labels derived from longitudinal cognitive assessments, then leveraged a semi-supervised framework to scale predictions to larger imputation cohorts lacking gold-standard cognition outcome labels. The algorithm robustly predicted the timing of first sustained cognitive worsening, with CDR outperforming MMSE and MoCA. For orthogonal validation, predicted time-to-worsening differentiated clinically meaningful subgroups defined by APOE-ε4 carrier status and knowledge graph-guided patient clustering.

**Implications of all the available evidence:** Scalable prediction of cognitive worsening from sparsely labeled EHR data could identify patients at higher risk of sustained cognitive worsening. In routine clinical practice, this could inform timely disease-modifying therapy initiation and care planning. Targeted enrichment of clinical trial populations with higher-risk patients could increase statistical power and reduce sample size requirements.

## INTRODUCTION

Alzheimer’s disease (AD) is a progressive neurodegenerative disorder affecting over 7.2 million people in the United States.^1–3^ The approval of disease-modifying therapies (DMTs) for early symptomatic AD and the expanding clinical trial pipeline necessitate reliable methods to evaluate real-world effectiveness and enrich clinical trial populations.^4–6^ Concurrently, as routine clinical practice introduces DMTs, there is a need for pragmatic and scalable monitoring of cognitive decline and treatment response using routinely collected clinical data, given that biomarkers (*e.g.,* fluid and neuroimaging markers) and cognitive assessments (*e.g.,* neuropsychological tests) that are rarely available in real-world settings.^7^

Cognitive assessments such as the Clinical Dementia Rating (CDR) scale, Mini-Mental State Examination (MMSE), and Montreal Cognitive Assessment (MoCA) are widely used in clinical trials and research registries to monitor disease progression by tracking cognitive decline, typically quantified as the change in score from baseline to a subsequent time point.^8–15^ In contrast to standardized administration in clinical trials, cognitive assessments in routine clinical practice are performed at irregular intervals, sparsely documented, and often administered in a subset of patients, with instrument choice varying across visits and settings. Two fundamental challenges constrain scalable monitoring of cognitive worsening. First, cognitive assessment documentation in electronic health records (EHRs) is sparse (3–33% across studies).^16–18^ Despite the potential of natural language processing and large language model-based technologies to extract cognitive scores from clinical narratives,^19–24^ longitudinal cognitive assessment data remain limited. Second, cognitive performance fluctuates across visits in AD patients due to individual-specific factors such as day-to-day symptom variation, comorbid conditions, and medication effects, as well as assessment-related factors such as test-retest reliability and practice effects from repeated administration.^25,26^ Worsening of a cognitive score at a single time point may reflect score fluctuation rather than true disease progression. This underscores the need for robust within-individual assessment of *sustained* cognitive worsening (*i.e.,* worsening that persists without improvement over consecutive visits), but the precise timing of such worsening is poorly documented in routine clinical practice.^27^

Sparsity and variability of longitudinal cognitive assessment data, coupled with the methodological challenge of ascertaining event timing from routine clinical documentation, have precluded the development of algorithms to predict sustained cognitive worsening over time. Rule-based methods largely rely on structured data and lack the flexibility to accurately extract event timing from unstructured clinical narratives,^19–24^ while traditional machine learning approaches require large-scale gold-standard labels for training.^28–30^ Semi-supervised and weakly supervised learning approaches could overcome label scarcity by leveraging unlabeled data or imperfect labels to augment sparse longitudinal labels, but predicting event timing introduces additional challenges, including modeling temporal dependencies, handling irregular visit intervals, and generating informative longitudinal surrogate features from unlabeled data.^30,31^

In this registry-linked EHR cohort study, we apply *label-efficient incident phenotyping from longitudinal EHR* (LATTE),^28^ a deep neural network algorithm optimized to identify clinical event timing from sparsely labeled longitudinal EHR data, to predict time-to-first sustained cognitive worsening across three complementary cognitive assessments (CDR, MMSE, and MoCA) in AD patients from a large healthcare system. We train LATTE on gold-standard cohorts with labels derived from longitudinal cognitive assessment scores, then leverage its semi-supervised learning framework to impute predictions across larger cohorts with insufficient cognitive assessment data. We evaluate clinical utility by examining whether predicted time-to-worsening differs across clinically meaningful patient subgroups.

## METHODS

### Ethics approval

The University of Pittsburgh Institutional Review Board approved the study protocol (STUDY21020153) and determined it to be exempt.

### Study population

We previously established an initial AD data mart comprising patients with ≥1 International Classification of Diseases (ICD) code for AD or related dementia (*e.g.,* ICD-9: 290.x, 294.2x, 331.0; ICD-10: F03.9x, G30.x) using inpatient and outpatient codified and narrative EHR data from the UPMC healthcare system (2011–2022).^32–34^ For the patient subset enrolled in the University of Pittsburgh Alzheimer’s Disease Research Center (ADRC) registry, we linked patient-level EHR data with registry elements (*e.g.*, AD diagnosis status, cognitive assessment scores). Our prior publications described the detailed EHR data structure, including narrative feature generation from clinical documentation using natural language processing. We identified patients with an AD diagnosis using either registry documentation in the ADRC subset or an unsupervised phenotyping algorithm (Knowledge-driven Online Multimodal Automated Phenotyping, KOMAP) for non-registry patients.^32–34^

### Cognitive assessment scores

We extracted and harmonized data for three commonly used and complementary cognitive assessments from both the EHR and ADRC registry: CDR, MMSE, and MoCA. CDR is typically administered in specialized AD care settings or clinical trials and not routinely documented in the EHR. CDR scores were available only from the ADRC registry. CDR is a dementia staging instrument that assesses six domains of cognitive and functional performance: memory, orientation, judgment and problem solving, community affairs, home and hobbies, and personal care.^8–12^ Each domain is rated on a five-point scale: 0 (none), 0.5 (questionable), 1 (mild), 2 (moderate), and 3 (severe). CDR-Sum of Boxes sums the individual domain scores, with total scores ranging from 0 to 18.^11^ CDR Global score is algorithmically derived from individual domain scores and rated on the same 5-point scale (0=none to 3=severe). Due to limited availability of CDR-Sum of Boxes in the ADRC registry, we used CDR Global for all analyses.

MMSE is a 30-point cognitive impairment screening tool, which assesses five domains of cognition: orientation, registration, attention and calculation, recall, and language and praxis.^13^ MoCA is a 30-point screening tool that assesses eight cognitive domains: visuospatial and executive function, naming, memory, attention, language, abstraction, delayed recall, and orientation.^14,15^ Compared to MMSE, MoCA enables more sensitive detection of mild cognitive impairment through broader assessment of visuospatial skills, executive function, and memory.

### First sustained cognitive worsening

From longitudinal cognitive assessment scores, we define sustained cognitive worsening by adapting minimum clinically important difference (MCID)-based thresholds for downstream analyses. MCID represents the smallest clinically meaningful difference in an outcome measure.^35^ Although MCID-based estimates vary by derivation cohort and follow-up duration, they reflect the available evidence for defining clinically meaningful cognitive change in AD populations.^27,36^ Seminal AD clinical trials (CLARITY AD, TRAILBLAZER-ALZ 2) operationalized pre-specified exploratory endpoints of time-to-worsening as any increase in CDR Global from randomization sustained over two consecutive scheduled visits, using MCID-based thresholds.^4,5,37–39^ Analogous MCID-based thresholds are established for MMSE and MoCA.^40,41^ We adapted these thresholds for use with EHR and ADRC registry cognitive assessment scores. For each cognitive assessment, we defined baseline cognitive impairment severity-specific thresholds for clinically meaningful worsening (hereafter, worsening thresholds). Baseline cognitive impairment severity for each assessment was categorized as normal (CDR 0–0.5, MMSE ≥24, MoCA ≥26, **Figure 1A**), mild (CDR 1, MMSE 19–23, MoCA 18–25), or moderate-to-severe (CDR 2–3, MMSE <18, MoCA <17).

**Figure 1.**
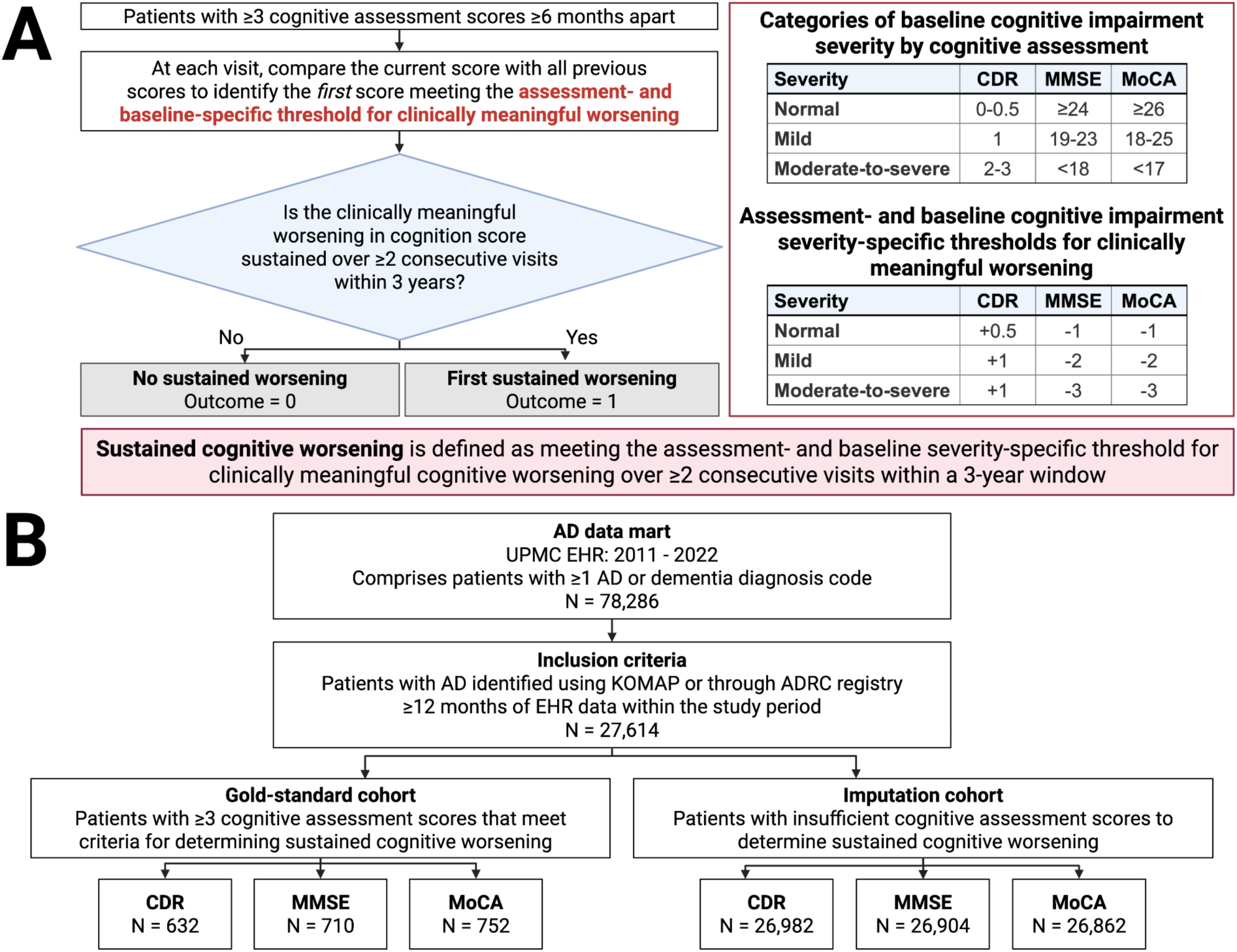
Outcome and cohort definition. (A) Stepwise approach to identifying the first sustained cognitive worsening from longitudinal cognitive assessments. We included patients with ≥3 cognitive assessment scores separated by ≥6 months. At each visit, we compared the current cognition score to all previous scores to identify the first worsening meeting or exceeding the assessment- and baseline cognitive impairment severity-specific threshold for clinically meaningful worsening.^4,5,37–41^ We defined *sustained cognitive worsening* as meeting this threshold at ≥2 consecutive visits within a 3-year window. We classified patients meeting these criteria as having “first sustained worsening” (outcome=1) and all others as “no sustained worsening” (outcome=0). (B) Cohort derivation. The flowchart illustrates cohort derivation from UPMC AD EHR data (2011–2022). From an initial data mart comprising patients with ≥1 diagnosis code for AD or dementia (N=78,286), we identified patients with an AD diagnosis (N=27,614) using an unsupervised phenotyping algorithm (KOMAP) or through linkage with the University of Pittsburgh Alzheimer’s Disease Research Center (ADRC) registry.^32,33^ We included patients with ≥12 months of EHR data within the study period. The gold-standard cohort for each cognitive assessment included AD patients with sufficient scores (≥3) to determine first sustained cognitive worsening (CDR, n=632; MMSE, n=710; MoCA, n=752). The corresponding imputation cohort comprised patients with insufficient scores (0–2) for whom the outcome required imputation (CDR, n=26,982; MMSE, n=26,904; MoCA, n=26,862). We evaluated model performance in the held-out subset of the gold-standard cohort, then applied the model to the imputation cohorts to scale the prediction of first sustained cognitive worsening.

We derived gold-standard labels of first sustained cognitive worsening among AD patients with ≥3 cognitive assessment scores and at least one pair of visits separated by ≥6 months (**Figure 1A**). At each visit, we compared the cognitive score to all prior scores to determine whether it met the worsening threshold (**Figure 1A, S-Table 1**). Visits meeting the worsening threshold were flagged for further confirmation. ***Sustained cognitive worsening*** was defined as two consecutive visits within a 3-year window that each met the worsening threshold, indicating persistence of decline rather than transient fluctuation. Patients meeting these criteria were classified as having “first sustained worsening” (outcome=1) and all others as “no sustained worsening” (outcome=0). Time-to-first sustained cognitive worsening was defined as the time in months from the first score in the sequence to the third score confirming sustained cognitive worsening.

**Table 1.** Baseline patient characteristics of gold-standard and imputation cohorts.

| Characteristic | Overall | CDR Global |  | MMSE |  | MoCA |  |
| --- | --- | --- | --- | --- | --- | --- | --- |
|  |  | Gold-standard<sup>a</sup> | Imputation<sup>b</sup> | Gold-standard<sup>a</sup> | Imputation<sup>b</sup> | Gold-standard<sup>a</sup> | Imputation<sup>b</sup> |
|  | N = 27,614 | N = 632 | N = 26,982 | N = 710 | N = 26,904 | N = 752 | N = 26,862 |
| Age at start of follow-up, Mean (SD) | 78.76 (9.53) | 70.12 (9.59) | 78.96 (9.44) | 71.37 (10.00) | 78.95 (9.44) | 69.97 (8.67) | 79.00 (9.44) |
| Women, N (%) | 17,943 (65%) | 358 (57%) | 17,585 (65%) | 410 (58%) | 17,533 (65%) | 443 (59%) | 17,500 (65%) |
| Race and ethnicity, N (%) |  |  |  |  |  |  |  |
| Non-Hispanic White | 25,226 (91%) | 565 (89%) | 24,661 (91%) | 637 (90%) | 24,589 (91%) | 706 (94%) | 24,520 (91%) |
| American Indian or Alaska Native | 13 (<1%) | 0 (0%) | 13 (<1%) | 0 (0%) | 13 (<1%) | 1 (<1%) | 12 (<1%) |
| Asian | 110 (<1%) | 6 (1%) | 104 (<1%) | 6 (1%) | 104 (<1%) | 6 (1%) | 104 (<1%) |
| Black | 2,171 (8%) | 58 (9%) | 2,113 (8%) | 63 (9%) | 2,108 (8%) | 35 (5%) | 2,136 (8%) |
| Hispanic or Latino | 89 (<1%) | 3 (<1%) | 86 (<1%) | 4 (1%) | 85 (<1%) | 4 (1%) | 85 (<1%) |
| Native Hawaiian or Other Pacific Islander | 4 (<1%) | 0 (0%) | 4 (<1%) | 0 (0%) | 4 (<1%) | 0 (0%) | 4 (<1%) |
| Other | 1 (<1%) | 0 (0%) | 1 (<1%) | 0 (0%) | 1 (<1%) | 0 (0%) | 1 (<1%) |
| Patients with APOE-ε4 data,<sup>c</sup> N | 1,028 | 628 | 400 | 619 | 409 | 376 | 652 |
| APOE-ε4 carrier, n (%) | 523 (51%) | 296 (47%) | 227 (57%) | 292 (47%) | 231 (56%) | 164 (44%) | 359 (55%) |
| APOE-ε4 non-carrier, n (%) | 505 (49%) | 332 (53%) | 173 (43%) | 327 (53%) | 178 (44%) | 212 (56%) | 293 (45%) |
| Patients eligible for clustering,<sup>d</sup> N | 14,247 | 155 | 14,092 | 192 | 14,055 | 301 | 13,946 |
| Cluster 1, n (%) | 3,900 (27%) | 40 (26%) | 3,860 (27%) | 50 (26%) | 3,850 (27%) | 103 (34%) | 3,797 (27%) |
| Cluster 2, n (%) | 10,347 (73%) | 115 (74%) | 10,232 (73%) | 142 (74%) | 10,205 (73%) | 198 (66%) | 10,149 (73%) |
| Death during follow-up, N (%) | 16,483 (60%) | 159 (25%) | 16,324 (60%) | 211 (30%) | 16,272 (60%) | 196 (26%) | 16,287 (61%) |

- Gold-standard cohorts included patients with sufficient cognitive assessment scores ( $\geq 3$ ) to determine first sustained cognitive worsening. - Imputation cohorts included patients with insufficient cognitive assessment scores (0-2) for whom the first sustained cognitive worsening outcome required imputation. - APOE-ε4* data were available in a subset of patients through linkage with the ADRC registry. - We excluded patients who experienced first sustained cognitive worsening or had <24 months of EHR data before AD diagnosis from the clustering analysis.
Abbreviations: CDR, Clinical Dementia Rating; MMSE, Mini-Mental State Examination; MoCA, Montreal Cognitive Assessment.

### Gold-standard vs. imputation cohort

We included UPMC AD patients with ≥12 months of codified and narrative EHR data within the study period (**Figure 1B**). The gold-standard cohort for each cognitive assessment included patients with sufficient scores (≥3) to determine first sustained cognitive worsening. The imputation cohort comprised patients with insufficient scores (0–2) for whom the outcome required imputation.

### Data for modeling

To prepare for longitudinal modeling, EHR data (2011–2022) were binned into consecutive, non-overlapping three-month intervals, with follow-up duration truncated at 9 years, equivalent to 36 three-month intervals (**Figure 2A**). Model input features (**Figure 2B**) included time-invariant demographics (*e.g.,* sex, race, and ethnicity) and time-varying features aggregated for each three-month interval (*e.g.,* age, comorbidities, healthcare utilization, and AD-related features). Gender was categorized as men and women. Due to the modest sample size of racial and ethnic minority groups within the UPMC catchment area, we dichotomized race and ethnicity as non-Hispanic White versus all other groups, including American Indian or Alaska Native, Asian, Black, Native Hawaiian or Pacific Islander, and others. Healthcare utilization was measured as the count of unique codified and narrative EHR features in each three-month interval. We quantified comorbidities using the Elixhauser Comorbidity Index, supplemented with AD-related comorbidities (*e.g.,* stroke, traumatic brain injury, and obstructive sleep apnea).^42,43^ We identified AD-related features from a multi-source knowledge graph^32^ based on cosine similarity to AD, with higher values indicating stronger feature association with AD. Additional inputs included gold-standard labels of observed first sustained cognitive worsening derived from cognitive assessment scores, and silver-standard labels of cumulative predicted probability of AD diagnosis status.^32,33^ We previously used multi-source knowledge-graph representation learning to select AD-related codified and narrative EHR features, then trained a phenotyping algorithm (KOMAP) on the covariance structure of these features to predict and identify patients with AD diagnosis. Here, we applied KOMAP cumulatively at each three-month interval to obtain a time-varying predicted probability of AD diagnosis. Finally, we included population-level AD-related feature embeddings to initialize the concept reweighting module.^32^

**Figure 2.**
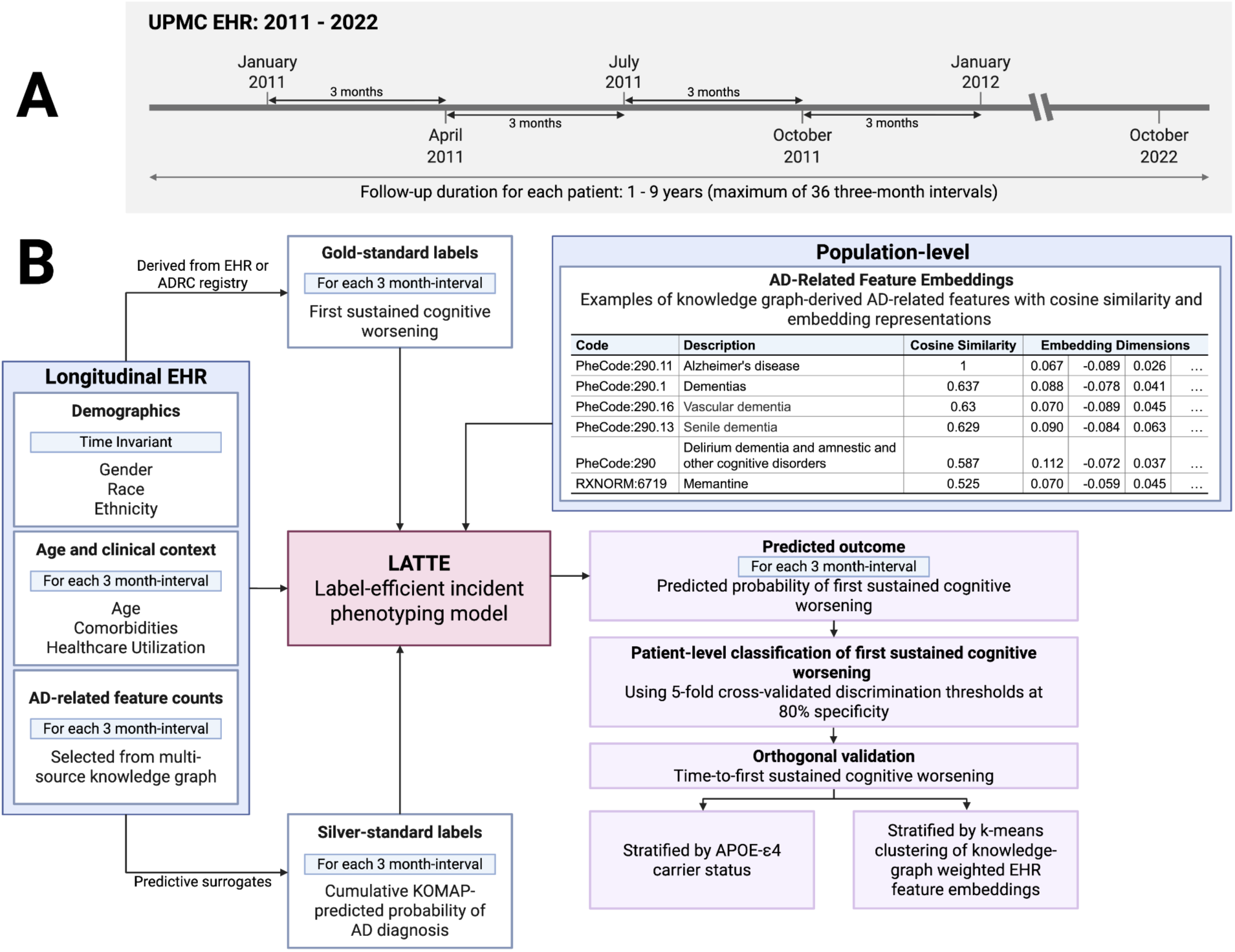
Study overview. **(A) Study timeline.** We binned longitudinal UPMC EHR data (2011–2022) into consecutive, non-overlapping three-month intervals (*e.g.,* January to April 2011, continuing until end of follow-up). For each patient, we truncated follow-up duration at 9 years, equivalent to 36 three-month intervals. **(B) LATTE model inputs and data structure.** Input features for LATTE included: (1) longitudinal patient-level EHR data with time-invariant (*e.g.,* race) and time-varying features aggregated for each three-month interval (*e.g.,* age, comorbidities, healthcare utilization, and AD-related features selected from a multi-source knowledge graph^32^); (2) gold-standard labels of first sustained cognitive worsening for each three-month interval; (3) silver-standard labels of cumulative KOMAP-predicted probability of AD diagnosis for each three-month interval;^32,33^ and (4) population-level AD-related feature embeddings derived from a multi-source knowledge graph.^32^ The table shows examples of AD-related features along with cosine similarity scores (higher values indicate stronger feature association with AD) and the first few embedding dimensions (embeddings are numerical representations that encode the clinical relationships between features and AD). LATTE uses semi-supervised learning to predict the probability of first sustained cognitive worsening for each three-month interval. We converted these interval-level predicted probabilities to patient-level classifications of first sustained cognitive worsening using a discrimination threshold. To demonstrate clinical utility, we performed orthogonal validation by examining whether time-to-first sustained cognitive worsening differentiates clinically meaningful patient subgroups defined by *APOE*-ε4 carrier status and k-means clustering of knowledge-graph-weighted EHR feature embeddings.

### Overview of modeling framework

To predict the timing of first sustained cognitive worsening, we applied **LATTE**, a deep neural network algorithm optimized for longitudinal phenotyping in data settings with sparse gold-standard labels.^28,30^ LATTE comprises four components: (1) a concept re-weighting module, which selects predictive EHR features related to the outcome using pre-trained semantic embeddings from a multi-source knowledge graph;^32^ (2) a visit attention network, which distinguishes informative visits from background noise; (3) a bidirectional gated recurrent unit (Bi-GRU) network, which models the sequential dependency among visits; and (4) incident predictors, which generate outcome predictions at each follow-up interval. To reduce reliance on longitudinal gold-standard labels, LATTE optimizes these four components in two stages. In unsupervised pre-training, silver-standard labels are constructed from predictive surrogates of the gold-standard outcome to learn longitudinal feature patterns and temporal dependencies inherent in the EHR data. In semi-supervised fine-tuning, model parameters are optimized separately using gold-standard and silver-standard labels to prevent noise in the silver-standard labels from degrading learning from gold-standard labels.

### Predicting the timing of first sustained cognitive worsening

We divided the gold-standard cohorts into training (80%) and test (20%) sets at the patient level, stratified by outcome. For each cognitive assessment (CDR, MMSE, MoCA), we trained a separate LATTE model on the respective gold-standard training set to predict the probability of first sustained cognitive worsening at each three-month interval and evaluated model performance on the corresponding held-out gold-standard test set. We then applied each model to the corresponding imputation cohort to extend predictions to patients with insufficient cognitive assessment data.

### Statistical Methods

#### Evaluating model performance

We evaluated model performance using four *longitudinal classification* metrics. First, average time-specific Area Under the Receiver Operating Characteristic Curve (**Average Time-AUC**) quantifies discrimination as the mean of the time-specific AUC computed at each three-month interval during follow-up. Each time-specific AUC reflects the model’s ability to distinguish patients who experience first sustained cognitive worsening at that interval (outcome=1) from those who do not (outcome=0). Higher average time-specific AUC values indicate better discrimination. Second, area between curves (**ABC**) quantifies calibration as the mean absolute difference between the observed and predicted cumulative incidence curves across all three-month intervals during follow-up.^28^ Lower ABC indicates better calibration. Third, **Brier score** measures overall predictive accuracy as the mean squared difference between the observed outcome and the predicted probability of first sustained cognitive worsening, averaged across all three-month intervals during follow-up. Lower Brier score indicates better performance. Finally, we assessed post-worsening survival time among patients predicted to experience first sustained cognitive worsening as the time in months from the predicted onset of first sustained cognitive worsening to death or end of follow-up. For each cognitive assessment, we performed two sensitivity analyses in the gold-standard cohorts, stratifying model performance by baseline cognitive impairment severity (*i.e.,* normal or mild) and by cognitive assessment data source (*i.e.,* ADRC, EHR, or both).

#### Classifying first sustained cognitive worsening

To convert interval-level predicted probabilities to patient-level classifications of first sustained cognitive worsening, we used 5-fold cross-validation (with 1,000 bootstrap replicates) within the gold-standard training sets to identify a discrimination threshold that achieved 80% specificity, maintaining reasonable sensitivity and positive predictive value (PPV) while minimizing false positives. We then applied this threshold to the gold-standard test sets and evaluated patient-level classification performance using AUC, sensitivity, PPV, and negative predictive value (NPV). Patients whose predicted probability exceeded the threshold at a given three-month interval were classified as experiencing first sustained cognitive worsening at that interval. For the imputation cohorts, predicted probabilities were recalibrated to match the observed outcome prevalence in the corresponding gold-standard cohort before applying the same discrimination threshold. Predicted time-to-first sustained cognitive worsening was defined as the time in months from the start of follow-up to the first three-month interval at which the predicted probability exceeded the discrimination threshold.

#### Assessing clinical utility

As orthogonal validation and to assess clinical utility, we examined whether the predicted timing of first sustained cognitive worsening differentiated clinically meaningful patient subgroups in the gold-standard, imputation, and combined cohorts. First, we compared the predicted outcome between *APOE*-ε4 carriers and non-carriers using unadjusted competing-risk Cox proportional hazards models accounting for death. This analysis was unadjusted because we hypothesized that key covariates (*e.g.,* age, comorbidities, healthcare utilization) mediate rather than confound the association between *APOE*-ε4 carrier status and cognitive worsening. Adjusting for variables on the causal pathway would underestimate the total effect.^44,45^

Second, we stratified patients into subgroups using k-means clustering based on their baseline EHR profiles (from the 24 months before the index date). For this analysis, we excluded patients who experienced first sustained cognitive worsening prior to the index date (first occurrence of AD diagnosis code) or who had <24 months of prior EHR data.^34^ We applied k-means clustering to patient embeddings generated by weighting the AD-related feature counts used as LATTE input features (in the two years prior to the index date) using population-level AD-related feature embeddings from a multi-source knowledge graph,^32^ adjusting for healthcare utilization. We then applied principal component analysis to reduce dimensionality, retaining components that explained 95% of the cumulative variance. We selected the optimal number of clusters using the silhouette method, which measures how well each patient fits into their assigned cluster relative to other clusters.^46^ We compared the predicted outcome among patient subgroups using unadjusted competing-risk Cox proportional hazards models, accounting for death as a competing event. We also examined the AD-related features distinguishing each cluster by regressing log-transformed feature counts against log-transformed healthcare utilization. We calculated feature importance as the difference in mean residuals between the two clusters, scaled by the standard deviation of each feature. Feature importance reflects the relative enrichment of each feature in one cluster versus the other, after accounting for variation in healthcare utilization.

#### Data availability

Anonymous summary-level registry data and EHR data will be publicly available. Patient-level data will not be publicly available because patient-level clinical data, whether de-identified or containing limited protected health information such as dates of clinical events (or even if anonymous due to the potential risk of re-identification), are universally subject to the rules and regulations of the healthcare system, which is affiliated with but not the same as the primary academic institution of the study investigators. De-identified data may be shared with qualified external researchers upon reasonable request to the corresponding authors and approval of the relevant Institutional Review Boards (IRBs), regulatory oversight bodies that hold custodianship of patient data within the healthcare system as well as appropriate Data Use Agreements (DUA) between institutions.

#### Software and code availability

Statistical analyses were conducted using Python (3.6.9) or R (version 4.4.1). Codes for KOMAP, LATTE, and project analyses are publicly available on Github.^28,32,47^

## RESULTS

### Patient characteristics

The entire cohort comprised 27,614 AD patients (65% women, 91% non-Hispanic White, mean [SD] age = 78.76 [9.53] years at follow-up start) (**Table 1**). Compared to imputation cohorts, gold-standard cohorts had a lower proportion of women (% women; gold-standard: CDR, 57%, MMSE, 58%, MoCA, 59%; imputation: CDR, 65%, MMSE, 65%, MoCA, 65%) and comprised younger patients (mean [SD] age; gold-standard: CDR, 70.12 [9.59], MMSE, 71.37 [10.00], MoCA, 69.97 [8.67]; imputation: CDR, 78.96 [9.44], MMSE, 78.95 [9.44], MoCA, 79.00 [9.44]). The proportion of racial and ethnic groups was largely consistent across cohorts (% non-Hispanic White; gold-standard: CDR, 89%, MMSE, 90%, MoCA, 94%; imputation: CDR, 91%, MMSE, 91%, MoCA, 91%). Among the 1,028 patients with available *APOE*-ε4 data, 523 (51%) were *APOE*-ε4 carriers, with higher carrier prevalence in the imputation cohorts than in the gold-standard cohorts (% *APOE*-ε4 carriers; gold-standard: CDR, 47%, MMSE, 47%, MoCA, 44%; imputation: CDR, 57%, MMSE, 56%, MoCA, 55%). During follow-up, 16,483 (60%) patients died, with lower mortality in the gold-standard cohorts than in the imputation cohorts (% death; gold-standard: CDR, 25%, MMSE, 30%, MoCA, 26%; imputation: CDR, 60%, MMSE, 60%, MoCA, 61%).

### Predicting timing of first sustained cognitive worsening

The gold-standard cohorts varied in sample size across cognitive assessment types: CDR, 632 patients (507 training, 125 test); MMSE, 710 patients (569 training, 141 test); MoCA, 752 patients (602 training, 150 test) (**S-Table 2, S-Table 3**). These gold-standard cohorts are not mutually exclusive, as patients with sufficient scores (≥3) for multiple assessments could appear in more than one cohort.

LATTE reasonably predicted the timing of first sustained cognitive worsening in the gold-standard test sets, with CDR outperforming MMSE and MoCA. Average-time AUC was highest for CDR, followed by MoCA and MMSE (CDR, 0.816; MMSE, 0.694; MoCA, 0.710) (**Figure 3**, **S-Table 4**, **S-Table 5**), with higher values during the initial phase of follow-up (**S-Figure 1**, **S-Figure 2**). ABC (CDR, 0.067; MMSE, 0.293; MoCA, 0.078) and Brier scores (CDR, 0.252; MMSE, 0.437; MoCA, 0.295) were lower for CDR and MoCA than for MMSE (**Figure 3**, **S-Table 5**).

**Figure 3.**
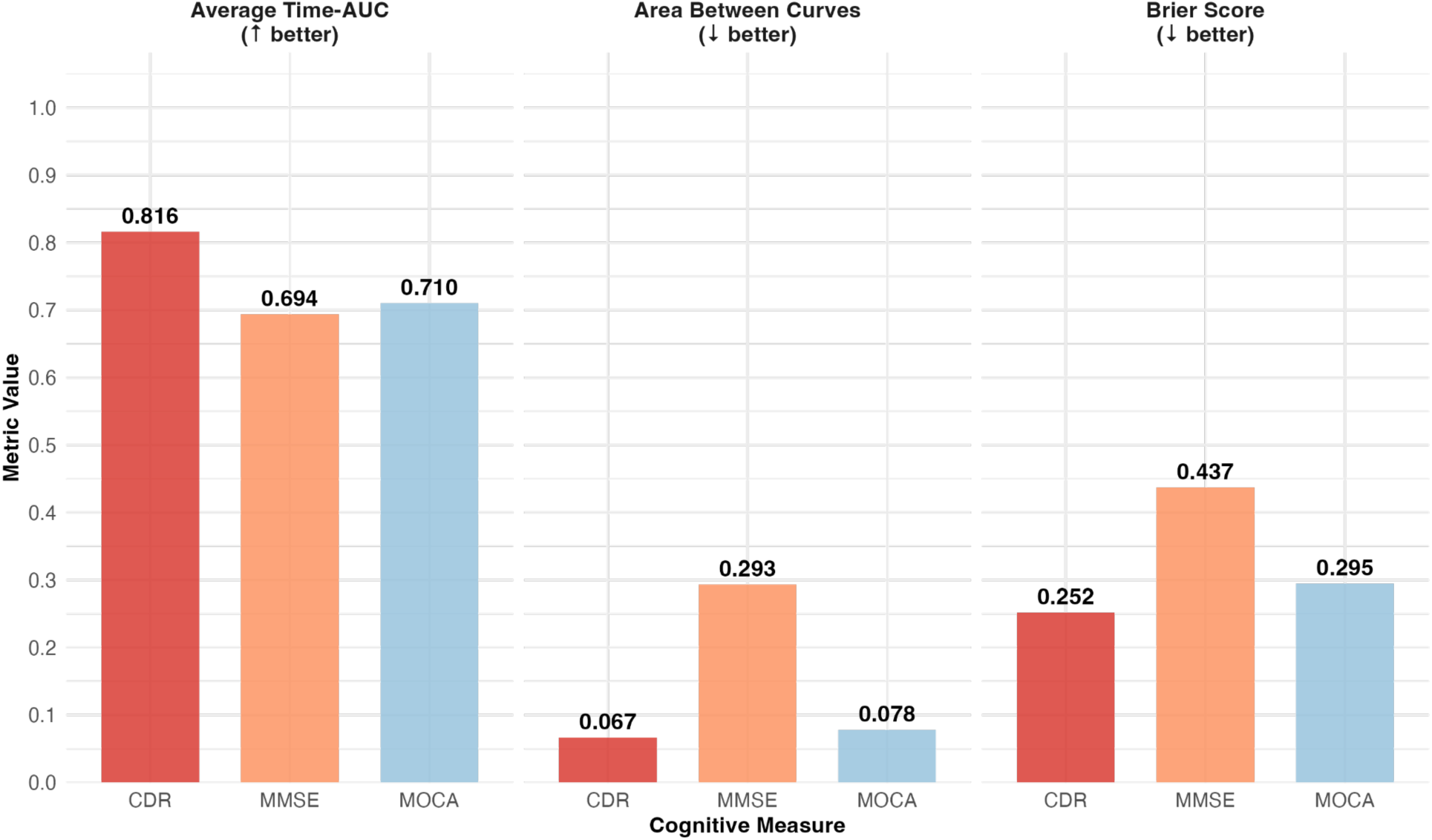
Performance of predicting first sustained cognitive worsening across cognitive assessments. We report longitudinal model performance metrics for CDR, MMSE, and MoCA scores. Average time-Area Under the Receiver Operating Characteristic Curve (Average Time-AUC) quantifies discrimination as the mean of the time-specific AUC computed at each three-month interval during follow-up, with higher values indicating better model discrimination. Area Between Curves (ABC) quantifies calibration as the mean absolute difference between the observed and predicted cumulative incidence curves across all three-month intervals during follow-up, with lower values indicating better model calibration.^28^ Brier score measures overall predictive accuracy as the mean squared difference between the observed outcome and predicted probability of first sustained cognitive worsening, averaged across all three-month intervals during follow-up, with lower values indicating better model performance.

Among patients predicted to experience first sustained cognitive worsening, MMSE had the longest post-worsening survival time, followed by MoCA, and CDR had the shortest (median [IQR] post-worsening survival, months; CDR, 42 [12, 96]; MMSE, 78 [39, 96]; MoCA: 54 [21, 102]) (**S-Table 5**). A long post-worsening survival time for MMSE indicates that predicted first sustained cognitive worsening occurred earliest in follow-up, whereas a short post-worsening survival time for CDR indicates later predicted onset.

### Sensitivity analyses

In the first sensitivity analysis, we report model performance stratified by baseline cognitive impairment severity. We included patients with normal or mild baseline scores and excluded the moderate-to-severe group due to small sample size (**S-Table 6**). Although combined models encompassing all baseline severity groups achieved the strongest overall performance, performance remained robust within the normal and mild groups, with some variation across cognitive assessments (**S-Figure 3**). In the second sensitivity analysis, we report model performance stratified by cognitive assessment data source. Cognitive assessment scores were predominantly derived from the ADRC registry, with additional MMSE and MoCA scores available from the EHR (**S-Table 7**). EHR-derived scores showed higher average-time AUC but poorer ABC and Brier scores compared to ADRC registry-derived scores for both MMSE and MoCA (**S-Figure 4**).

**Figure 4.**
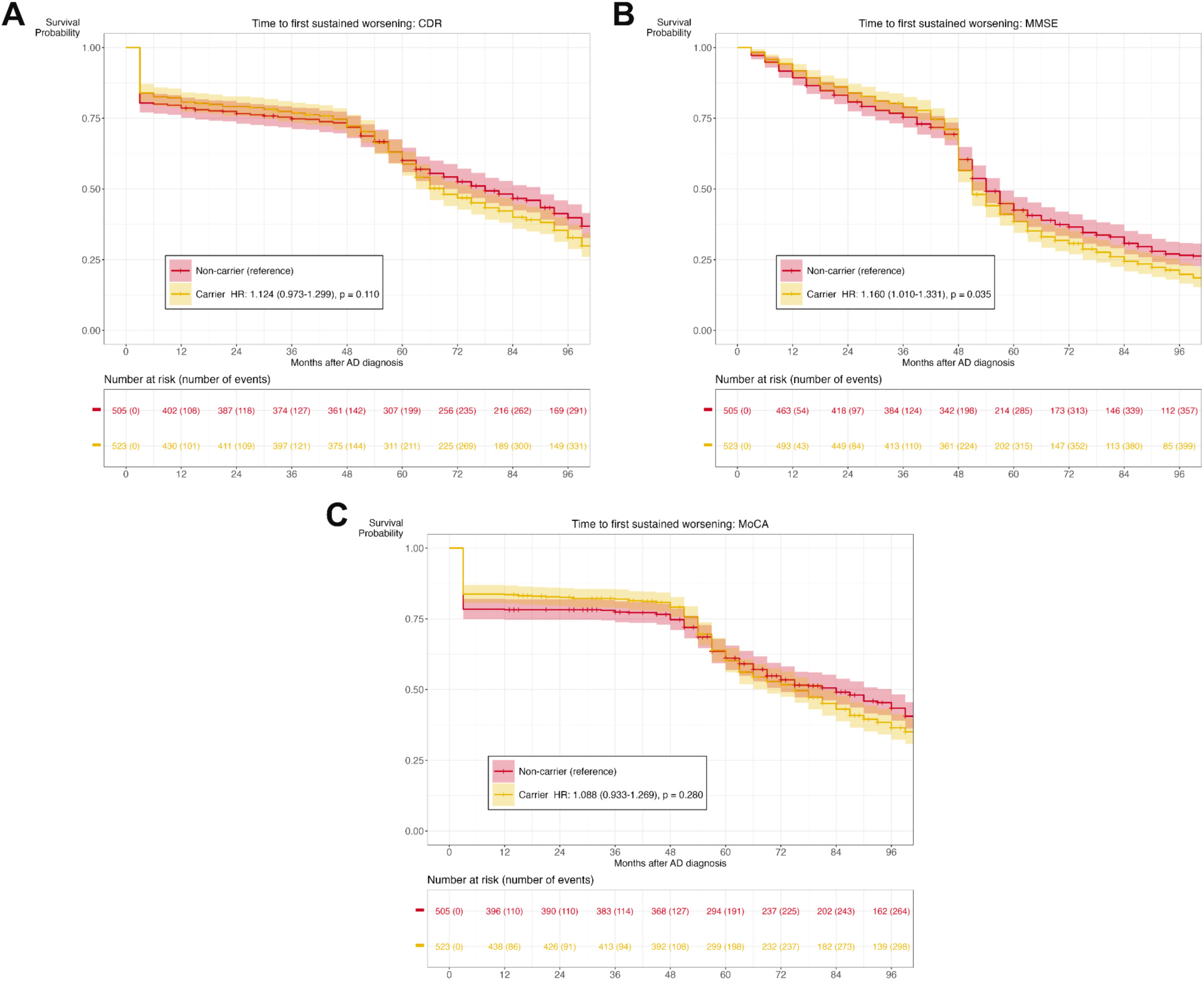
Time-to-first sustained cognitive worsening stratified by *APOE*-ε4 carrier status. We estimated time-to-first sustained cognitive worsening across all cognitive assessments using unadjusted, competing-risk Cox proportional hazards models, accounting for death as a competing event. Solid lines represent survival probability. Shaded regions represent 95% confidence intervals. The reference group for *APOE*-ε4 carrier status was non-carrier. Combined cohorts included gold-standard and imputed patients. **(A) CDR Global, combined cohort** (n=1028; 679 events). **(B) MMSE, combined cohort** (n=1028; 764 events). **(C) MoCA, combined cohort** (n=1028; 595 events).

### Classifying patients reaching first sustained cognitive worsening

We then report LATTE performance in classifying patients reaching first sustained cognitive worsening (**S-Table 8**). At the patient level, LATTE achieved the strongest performance for CDR (metric [95% CI]: AUC 0.831 [0.811–0.852], sensitivity 0.750 [0.703–0.793], PPV 0.526 [0.480–0.573], NPV 0.916 [0.901–0.930]), followed by MoCA (AUC 0.770 [0.747–0.791], sensitivity 0.620 [0.581–0.661], PPV 0.523 [0.484–0.563], NPV 0.857 [0.840–0.872]). The high NPV for both CDR and MoCA suggests robust performance in ruling out first sustained cognitive worsening, whereas the lower PPV likely reflects class imbalance in the gold-standard cohorts (outcome prevalence: CDR, 22%; MoCA, 26%) (**S-Table 2**). MMSE achieved lower AUC (0.738 [0.718–0.758]), sensitivity (0.533 [0.496–0.568]), and NPV (0.661 [0.640–0.684]), but higher PPV (0.701 [0.668–0.735]), consistent with its higher outcome prevalence (outcome prevalence: MMSE, 48%) (**S-Table 2**).

### Time-to-first sustained cognitive worsening stratified by *APOE*-ε4 carrier status

In unadjusted competing-risk Cox proportional hazards models of time-to-first sustained cognitive worsening, accounting for death as a competing event, *APOE*-ε4 carriers in the overall cohort had shorter predicted time-to-worsening than non-carriers on the MMSE (HR [95% CI]: 1.160 [1.010–1.331], *p*=.035) (**Figure 4**), but not on the CDR (1.124 [0.973–1.299], *p*=.110) or MoCA (1.088 [0.933–1.269], *p*=.280). In gold-standard cohorts, no significant difference was observed on any cognitive assessment (CDR, 1.116 [0.944–1.318], *p*=.200; MMSE, 1.097 [0.905–1.329], *p*=.350; MoCA, 1.168 [0.980–1.392], *p*=.082) (**S-Figure 5**). In imputation cohorts, however, *APOE*-ε4 carriers had shorter predicted time-to-worsening across all three cognitive assessments (CDR, 1.376 [1.048–1.806], *p*=.022; MMSE, 1.241 [1.030–1.496], *p*=.023; MoCA, 1.336 [1.054–1.693], *p*=.017) (**S-Figure 6**).

**Figure 5.**
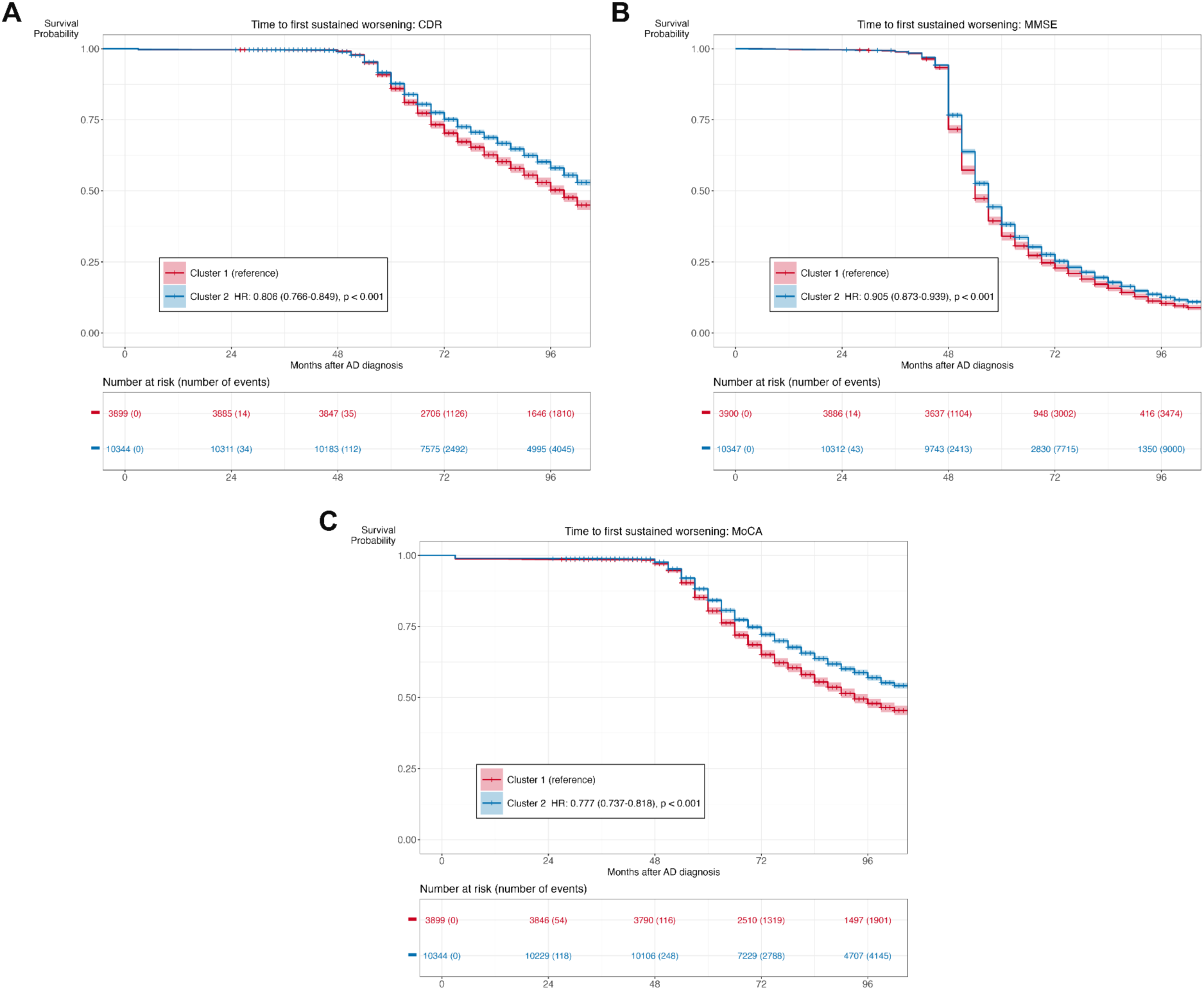
Time-to-first sustained cognitive worsening stratified by baseline patient clustering. We excluded patients with first cognitive worsening or <24 months of EHR data prior to AD diagnosis. We clustered the remaining patients into two subgroups using k-means clustering (k=2) of knowledge-graph-weighted EHR feature embeddings, adjusted for healthcare utilization (n = 14,247 AD patients; Cluster 1: n = 3,900; Cluster 2: n = 10,347). We estimated time-to-first sustained cognitive worsening using unadjusted competing-risk Cox proportional hazards models accounting for death as a competing event. Solid lines represent survival probability. Shaded regions represent 95% confidence intervals. Cluster 1 served as the reference group. Combined cohorts included gold-standard and imputed patients. **(A) CDR Global, combined cohort** (n=14,247; 6,404 events). **(B) MMSE, combined cohort** (n=14,247; 12,686 events). **(C) MoCA, combined cohort** (n=14,247; 6,326 events).

### Time-to-first sustained cognitive worsening stratified by baseline patient clustering

After excluding patients who experienced first sustained cognitive worsening prior to the index date or had <24 months of prior EHR data, 14,247 of 27,614 patients were eligible for stratification. K-means clustering according to baseline EHR profile with adjustment for healthcare utilization identified two subgroups (Cluster 1: 3,900; Cluster 2: 10,347) (**Table 1, S-Figure 7**). In unadjusted competing-risk Cox proportional hazards models of time-to-first sustained cognitive worsening, accounting for death as a competing event, Cluster 2 had longer predicted time-to-worsening than Cluster 1 across all three cognitive assessments in the combined cohort (HR [95% CI]; CDR, 0.806 [0.766–0.849], *p*<.001; MMSE, 0.905 [0.873–0.939], *p*<.001; MoCA, 0.777 [0.737–0.818], *p*<.001) (**Figure 5**). In the gold-standard cohorts, Cluster 2 had longer predicted time-to-worsening for CDR, but no significant difference was observed for MMSE or MoCA (CDR, 0.729 [0.531–1.000], *p*=.050; MMSE, 0.756 [0.551–1.038], *p*=.084; MoCA, 1.060 [0.869–1.293], *p*=.570) (**S-Figure 8**). Imputation cohort findings were consistent with those of the overall cohort (CDR, 0.805 [0.764–0.849], *p*<.001; MMSE, 0.908 [0.875–0.941], *p*<.001; MoCA, 0.777 [0.737–0.820], *p*<.001) (**S-Figure 9**). We examined features characteristic of each cluster (**S-Figure 10**). Cluster 1 was characterized by clinical factors indicative of high disease burden, including dementia-related diagnoses (*e.g.,* dementia, delirium, and amnestic and other cognitive disorders) and prescriptions (*e.g.,* donepezil), while Cluster 2 was characterized by comorbidity-related medication prescriptions (*e.g.,* amlodipine, tamsulosin).

## DISCUSSION

This study demonstrates the feasibility of predicting the timing of first sustained cognitive worsening from EHR data across three complementary cognitive assessments (CDR, MMSE, and MoCA) using LATTE, a deep neural network optimized for identifying clinical event timing from sparse longitudinal labels. Leveraging LATTE’s semi-supervised learning framework, we scaled predictions from gold-standard cohorts with sparse first sustained cognitive worsening labels to larger imputation cohorts with insufficient longitudinal cognitive assessment labels. Predicted time-to-worsening differentiated clinically meaningful patient subgroups: *APOE*-ε4 carrier status and patient subgroup clustering each independently predicted time-to-worsening.

LATTE demonstrated reasonable longitudinal predictive performance across all three assessments, with CDR prediction outperforming MMSE and MoCA for several reasons. First, CDR measures both cognitive and functional performance, whereas MMSE and MoCA assess largely cognitive domains.^8–15^ Functional decline may lag behind cognitive decline, which likely contributes to the later predicted onset of sustained worsening and shorter post-worsening survival observed for CDR. Second, we used CDR Global, which is less sensitive for tracking cognitive change than CDR-Sum of Boxes.^11^ Limited by few CDR-Sum of Boxes scores, using CDR Global scores may have simplified the prediction task and improved performance. Third, we obtained CDR scores exclusively through an EHR-linked ADRC registry whose participants likely have more reliable longitudinal trajectories than those from combined EHR and ADRC data used for MMSE and MoCA. While this interpretation is supported by greater variability in model performance for EHR-derived scores compared to ADRC registry-derived scores for MMSE and MoCA, we could not assess this for CDR because CDR scores were only available from ADRC.

Patient-level classification performance largely mirrored longitudinal predictive performance: CDR achieved the highest AUC, followed by MoCA and MMSE. CDR and MoCA had higher NPV than MMSE, while MMSE had the highest PPV. This is an expected finding given that MMSE had the highest outcome prevalence. High outcome prevalence increases PPV while decreasing NPV at any fixed specificity threshold. Since PPV and NPV are prevalence-dependent, these metrics should be interpreted with caution when predictions are scaled to larger cohorts with differing outcome prevalence. AUC, sensitivity, and specificity provide more stable measures of performance across deployment settings. Collectively, these findings suggest that the optimal cognitive assessment for clinical prediction depends on whether minimizing false positives or false negatives is the clinical priority.

We evaluated the clinical utility of the predicted first sustained cognitive worsening outcome through two complementary approaches. First, we examined whether the predicted outcome differed by *APOE*-ε4 carrier status. *APOE*-ε4 is the strongest genetic risk factor for AD, with carriers experiencing earlier age of onset and faster cognitive decline than non-carriers.^44,48^ Consistent with this, *APOE*-ε4 carriers had shorter predicted time-to-worsening across all three assessments in imputation cohorts, though not in gold-standard cohorts. This difference may reflect differential carrier prevalence between gold-standard and imputation cohorts and insufficient statistical power. Second, we stratified AD patients into two subgroups using k-means clustering of knowledge-graph-weighted baseline EHR feature embeddings. Cluster 2 had longer predicted time-to-worsening than Cluster 1 across all assessments in the overall and imputation cohorts. As with the *APOE*-ε4 analyses, inconsistent findings in the gold-standard cohorts likely reflect insufficient statistical power rather than a true absence of subgroup differences. Together, these findings indicate that the predicted outcome differentiates clinically meaningful patient subgroups and that predictions scale reliably from smaller gold-standard cohorts to larger imputation cohorts.

This study has several strengths. First, we developed a clinically grounded definition of first sustained cognitive worsening by adapting MCID-based thresholds for longitudinal EHR data. Requiring sustainment over consecutive visits reduces misclassification of transient cognitive fluctuations as true cognitive worsening. This approach provides an individualized, clinically interpretable measure of disease progression by evaluating cognitive worsening relative to each patient’s own baseline, offering an advantage over group-level predictions that can mask individual variability.^27^ Second, to our knowledge, this is the first study to predict the timing of first sustained cognitive worsening from longitudinal EHR data. This prediction task is challenging because the precise timing of first cognitive worsening and its sustainment are inherently difficult to ascertain from routine clinical documentation. To complement sparse gold-standard labels of first sustained cognitive worsening, we leveraged cumulative KOMAP-predicted probability of AD diagnosis as a silver-standard label for unsupervised pre-training, providing a dense longitudinal signal that enables LATTE to learn temporal patterns necessary for estimating clinical event timing.^28,32,33^ Although prior LATTE applications include predicting disease activity in other chronic neurological diseases (*e.g.,* acute relapse detection in multiple sclerosis),^28,30,49^ our application extends this method to predict a longitudinal outcome whose precise timing is poorly documented in routine clinical practice. Third, we demonstrate the feasibility of scaling predictions from smaller gold-standard cohorts to larger imputation cohorts. This addresses a key bottleneck, since most AD patients lack longitudinal cognitive assessments but represent the broader clinical population that could benefit from scalable monitoring of cognitive worsening. Finally, beyond evaluating model performance, we show that predicted outcomes are clinically meaningful and biologically plausible, as demonstrated by patient stratification strategies with established differences in AD risk and progression.

Our study also has several limitations. First, our definition of first sustained cognitive worsening (*i.e.,* decline persisting over ≥2 consecutive visits within a 3-year window) identifies the initial occurrence of sustained cognitive worsening. In future studies, we will characterize subsequent cognitive trajectories, including further worsening, stabilization, or potential recovery beyond the pre-defined sustainment window. Second, gold-standard cohorts predominantly comprised EHR-linked ADRC registry patients who were younger, had lower mortality, and were followed more regularly than imputation cohorts. This potential selection bias may limit the generalizability of scaling predictions to the broader AD population. Third, *APOE*-ε4 data were available only in a subset of patients through ADRC registry linkage, and differences in carrier prevalence between gold-standard and imputation cohorts may limit the interpretability of analyses stratified by *APOE*-ε4 status. Fourth, we applied k-means clustering to stratify patients at AD diagnosis based on 24 months of pre-index data. We chose k-means clustering to prioritize simplicity and interpretability, as the primary goal was to evaluate the clinical utility of the predicted outcomes rather than to optimize patient stratification.^34^ Although stratifying patients based on 24 months of pre-index data balances sample size and follow-up duration, this approach may not fully capture baseline patient heterogeneity, given that AD pathology often precedes diagnosis by decades.^50^ Finally, this study used data from a single healthcare system, though the study population included patients from academic and community practices across a wide catchment area. Prior work demonstrated the cross-institution portability of LATTE.^28^ Nevertheless, external validation in larger and more diverse cohorts will improve the generalizability of this AD application.

## CONCLUSION

In summary, LATTE reasonably predicted the timing of first sustained cognitive worsening from longitudinal EHR data and sparse labels across three complementary cognitive assessments. By leveraging a semi-supervised learning framework, LATTE enables scalable outcome ascertainment for the broader AD population where longitudinal cognitive assessment data are unavailable. Differentiation of clinically meaningful patient subgroups supports the clinical utility of these predictions. Predicting the timing of first sustained cognitive worsening in AD populations could have broad clinical and research applications. In routine clinical practice, early identification of patients at higher risk of sustained cognitive worsening could inform timely DMT initiation and care planning. Targeted enrichment of clinical trial populations with patients at higher risk of cognitive decline could increase statistical power to detect treatment effects while reducing sample size requirements, trial duration, and the cost of therapeutic development. Future directions include examining modifiable drivers of first sustained cognitive worsening, integrating additional data modalities (*e.g.,* fluid and neuroimaging biomarkers) to improve prediction, and externally validating findings in larger, more diverse cohorts.

## ACKNOWLEDGEMENTS

This study was supported by the National Institutes of Health under award numbers R01 NS098023 and R01 NS124882 from the National Institute of General Medical Sciences. The content is solely the responsibility of the authors and does not necessarily represent the official views of the National Institutes of Health.

## AUTHOR CONTRIBUTIONS

Shruthi Venkatesh, Sinian Zhang, Wen Zhu, Michele Morris, Rocco Mercurio, Sarah B. Berman, Hansruedi Mathys, Abby L. Olsen, C. Elizabeth Shaaban, Shyam Visweswaran, Oscar L. Lopez, Tianxi Cai, Jue Hou, and Zongqi Xia contributed to the design and conceptualization of the study. Shruthi Venkatesh and Sinian Zhang contributed equally as co-first authors to data analysis and manuscript writing. Wen Zhu, Michele Morris, Rocco Mercurio, Sarah B. Berman, Hansruedi Mathys, Abby L. Olsen, C. Elizabeth Shaaban, Shyam Visweswaran, Oscar L. Lopez, Tianxi Cai, Jue Hou, and Zongqi Xia contributed to data acquisition and manuscript writing. Jue Hou and Zongqi Xia contributed equally as co-senior authors and jointly supervised this work. All authors have reviewed and approved the final manuscript.

