## Supplementary Table 1-8 and Supplementary Figure 1-10 for "Predicting the timing of first sustained cognitive worsening in Alzheimer’s disease using real-world clinical data and machine learning"

### SUPPLEMENTARY MATERIAL

#### S-Table 1

Representative examples illustrating the identification of first sustained cognitive worsening. [a]

| Baseline cognitive impairment severity [b] | Cognitive assessment | Period | 1 | 3 | 5 | 8 | 11 | 13 | 16 | 20 | 25 |
|---|---|---|---|---|---|---|---|---|---|---|---|
| Normal | CDR Global | Cognition score | 0 | - | 0.5 | 0 | 0.5 | - | 0.5 | 0.5 | - |
| | | Outcome [c] | 0 | - | 0 | 0 | 0 | - | 1 | 1 | - |
| | MMSE | Cognition score | 27 | - | 28 | 26 | - | 24 | - | - | 23 |
| | | Outcome | 0 | - | 0 | 0 | - | 1 | - | - | 1 |
| | MoCA | Cognition score | - | - | 28 | 27 | - | 26 | - | 25 | - |
| | | Outcome | - | - | 0 | 0 | - | 1 | - | 1 | - |
| Mild | CDR Global | Cognition score | - | 1 | - | - | 1 | - | 2 | 2 | - |
| | | Outcome | - | 0 | - | - | 0 | - | 0 | 1 | - |
| | MMSE | Cognition score | 21 | 23 | - | 23 | - | 20 | 20 | - | - |
| | | Outcome | 0 | 0 | - | 0 | - | 0 | 1 | - | - |
| | MoCA | Cognition score | - | - | 23 | 24 | - | 24 | - | 22 | 21 |
| | | Outcome | - | - | 0 | 0 | - | 0 | - | 0 | 1 |
| Moderate-to-severe | CDR Global | Cognition score | - | 2 | - | - | 2 | - | 3 | 3 | 3 |
| | | Outcome | - | 0 | - | - | 0 | - | 0 | 1 | 1 |
| | MMSE | Cognition score | - | - | 17 | - | 16 | - | 12 | 11 | - |
| | | Outcome | - | - | 0 | - | 0 | - | 0 | 1 | - |
| | MoCA | Cognition score | 16 | 16 | - | - | 11 | 10 | - | 9 | - |
| | | Outcome | 0 | 0 | - | - | 0 | 1 | - | 1 | - |

a. For patients with ≥3 cognitive assessment scores each separated by ≥6 months, we compared the cognition score at each visit to all previous scores to identify the first score meeting or exceeding the assessment- and baseline cognitive impairment severity-specific threshold for clinically meaningful worsening.[4,5,37–41] We show hypothetical examples of cognitive score trajectories and first sustained cognitive worsening outcome classification.

b. We defined baseline as the first score in the sustained cognitive worsening endpoint. We categorized baseline cognitive impairment severity as normal, mild, or moderate-to-severe, and set thresholds for clinically meaningful worsening for each category: CDR Global (Normal +0.5, Mild AD +1, Moderate-to-Severe AD +1), MMSE (Normal -1, Mild AD -2, Moderate-to-Severe AD -3), and MoCA (Normal -1, Mild AD -2, Moderate-to-Severe AD -3).

c. We defined sustained cognitive worsening as reaching this threshold at ≥2 consecutive visits within a 3-year window. We classified patients meeting these criteria as "first sustained worsening" (outcome=1) and all others as “no sustained worsening” (outcome=0). After a patient reached first sustained worsening, we did not evaluate subsequent cognitive trajectories (*i.e.,* outcome=1 is retained at all subsequent visits).

Abbreviations: CDR, Clinical Dementia Rating; MMSE, Mini-Mental State Examination; MoCA, Montreal Cognitive Assessment

**S-Table 2**

Gold-standard cohort split for training and test.

| Cognitive assessment | Number of patients, N | Training, n [a] | Training Y=1 (%) [b] | Test, n [a] | Test Y=1 (%) [b] |
|---|---|---|---|---|---|
| CDR Global | 632 | 507 | 21.3 | 125 | 22.8 |
| MMSE | 710 | 569 | 48.5 | 141 | 46.8 |
| MoCA | 752 | 602 | 22.4 | 150 | 26.0 |

a. We divided patients in the gold-standard cohorts into training (80%) and test (20%) sets, stratified by the first sustained cognitive worsening outcome. We report patient counts for the training and test sets
b. We also report the proportion of patients experiencing first sustained cognitive worsening (Y=1).

Abbreviations: CDR, Clinical Dementia Rating; MMSE, Mini-Mental State Examination; MoCA, Montreal Cognitive Assessment

**S-Table 3**

Observed trajectories of first sustained cognitive worsening in gold-standard cohorts.

| Cognitive assessment | Number of patients, N [a] | First sustained cognitive worsening, n (%) [b] | Number of scores per patient, mean (SD) [c] | Duration between scores in months, mean (SD) [d] |
|---|---|---|---|---|
| CDR Global | 632 | 171 (27.1%) | 5.6 (4.2) | 13.0 (4.3) |
| MMSE | 710 | 411 (57.9%) | 5.6 (4.3) | 13.0 (4.4) |
| MoCA | 752 | 335 (44.5%) | 5.0 (3.5) | 11.2 (4.9) |

a. For patients in the gold-standard cohorts, we derived observed trajectories of first sustained cognitive worsening to serve as the gold-standard labeled outcome. We report:
b. The number and proportion of patients experiencing first sustained cognitive worsening;
c. The average number of scores per patient during follow-up;
d. The average duration between consecutive cognitive assessments in months.

Abbreviations: CDR, Clinical Dementia Rating; MMSE, Mini-Mental State Examination; MoCA, Montreal Cognitive Assessment

**S-Table 4**

Predicted trajectories of sustained cognitive worsening in gold-standard and imputation cohorts. [a]

| Cohort | Cognitive assessment | Number of patients, N | First sustained cognitive worsening, n (%) [b] | Number of scores per patient, mean (SD) [c] | Duration between scores in months, mean (SD) [d] |
|---|---|---|---|---|---|
| Gold-standard [e] | CDR Global | 632 | 471 (74.5%) | 29.5 (8.6) | 3.1 (0.2) |
| | MMSE | 710 | 481 (67.7%) | 31.1 (7.3) | 3.2 (0.6) |
| | MoCA | 752 | 659 (87.6%) | 31.7 (6.3) | 3.1 (0.5) |
| Imputation [f] | CDR Global | 26,982 | 16,373 (60.7%) | 33.7 (1.3) | 3.2 (0.1) |
| | MMSE | 26,904 | 25,044 (93.1%) | 33.7 (1.3) | 3.2 (0.1) |
| | MoCA | 26,862 | 16,007 (59.6%) | 33.7 (1.3) | 3.2 (0.1) |

a. Patients were classified as experiencing first sustained cognitive worsening if their predicted probability of first sustained cognitive worsening at any three-month interval exceeded the discrimination threshold. We report:
b. The number and proportion of patients experiencing predicted first sustained cognitive worsening (outcome=1);
c. The average number of cognitive assessment scores per patient;
d. The average duration between consecutive cognitive assessments in months.
e. For gold-standard cohorts, the discrimination threshold was derived from 5-fold cross-validation at 80% specificity in the training set.
f. For imputation cohorts, predicted probabilities were recalibrated to match the observed prevalence of first sustained cognitive worsening in the corresponding gold-standard cohort before applying the discrimination threshold.

Abbreviations: CDR, Clinical Dementia Rating; MMSE, Mini-Mental State Examination; MoCA, Montreal Cognitive Assessment

**S-Table 5**

Longitudinal model performance metrics.

| Cognitive assessment [a] | Number of patients, N | Average-time AUC [b] | Area between curves [c] | Brier score [d] | Post-worsening survival time in months, median [IQR] [e] |
|---|---|---|---|---|---|
| CDR Global | 632 | 0.816 | 0.067 | 0.252 | 42 [12, 96] |
| MMSE | 710 | 0.694 | 0.293 | 0.437 | 78 [39, 96] |
| MoCA | 752 | 0.710 | 0.078 | 0.295 | 54 [21, 102] |

a. We report longitudinal model performance for each cognitive assessment type in gold-standard test sets using three measures:
b. Average time-AUC (mean time-specific area under the receiver operating characteristic curve across all three-month intervals during follow-up, where higher values indicate better model discrimination);
c. Area between curves (mean absolute difference between the observed and predicted cumulative incidence curves across follow-up, where lower values indicate better model calibration);
d. Brier score (mean squared difference between the observed gold-standard outcome and predicted probability of first sustained cognitive worsening at each three-month interval, where lower values indicate better model predictive accuracy).
e. We also report post-worsening survival time among patients predicted to experience first sustained cognitive worsening as the time in months from the predicted onset of first sustained cognitive worsening to death or end of follow-up.

<u>Abbreviations</u>: AUC, Area Under the Receiver Operating Characteristic Curve; CDR, Clinical Dementia Rating; MMSE, Mini-Mental State Examination; MoCA, Montreal Cognitive Assessment

**S-Table 6**

Observed trajectories of first sustained cognitive worsening in gold-standard cohorts, stratified by baseline cognitive impairment severity. [a]

| Cognitive assessment | Baseline cognitive impairment severity [b] | Number of patients, N | First sustained worsening, n (%) [c] | Number of scores per patient, mean (SD) [d] | Duration between scores in months, mean (SD) [e] |
|---|---|---|---|---|---|
| CDR Global | Normal | 493 | 118 (23.9%) | 5.5 (3.5) | 13.4 (4.5) |
| | Mild | 116 | 50 (43.1%) | 5.6 (4.9) | 11.6 (2.9) |
| | Moderate-to-severe | 23 | 3 (13%) | 8.8 (9.9) | 10.4 (4.1) |
| MMSE | Normal | 533 | 304 (57%) | 5.6 (3.9) | 13.4 (4.7) |
| | Mild | 131 | 94 (71.8%) | 5.4 (4.9) | 11.9 (2.9) |
| | Moderate-to-severe | 46 | 13 (28.3%) | 5.6 (6.5) | 11.3 (3.4) |
| MoCA | Normal | 158 | 58 (36.7%) | 4.6 (3.1) | 13.8 (4.5) |
| | Mild | 355 | 187 (52.7%) | 5.3 (3.5) | 11.0 (4.9) |
| | Moderate-to-severe | 239 | 90 (37.7%) | 4.8 (3.7) | 9.9 (4.4) |

a. We derived first sustained cognitive worsening trajectories for each baseline cognitive impairment severity category in gold-standard cohorts.
b. Baseline was defined as the first cognitive assessment score in the sustained cognitive worsening endpoint. Baseline cognitive impairment severity was categorized as normal, mild, or moderate-to-severe. For each severity category and cognitive assessment, we report:
c. The number and proportion of patients experiencing first sustained cognitive worsening;
d. The average number of scores per patient;
e. The average duration between consecutive cognitive assessments in months.

Abbreviations: AUC, Area Under the Receiver Operating Characteristic Curve; CDR, Clinical Dementia Rating; MMSE, Mini-Mental State Examination; MoCA, Montreal Cognitive Assessment

**S-Table 7**

Observed trajectories of first sustained cognitive worsening in gold-standard cohorts, stratified by cognitive assessment data source. [a]

| Data source [b] | Cognitive assessment | Number of patients, N | First sustained worsening, n (%) [c] | Number of scores per patient, mean (SD) [d] | Duration between scores in months, mean (SD) [e] |
|---|---|---|---|---|---|
| Combined | MMSE | 710 | 411 (57.9%) | 5.6 (4.3) | 13.0 (4.4) |
| | MoCA | 752 | 335 (44.5%) | 5.0 (3.5) | 11.2 (4.9) |
| ADRC | CDR Global [f] | 632 | 171 (27.1%) | 5.6 (4.2) | 13.0 (4.3) |
| | MMSE | 617 | 342 (55.4%) | 5.6 (4.2) | 13.0 (4.3) |
| | MoCA | 368 | 145 (39.4%) | 4.5 (3.3) | 13.2 (3.4) |
| EHR | MMSE | 90 | 66 (73.3%) | 5.6 (5.1) | 13.2 (4.7) |
| | MoCA | 381 | 183 (48%) | 5.5 (3.7) | 9.3 (5.1) |

a. We derived first sustained cognitive worsening trajectories for each cognitive assessment data source in gold-standard cohorts.
b. Data sources include ADRC registry data, EHR data, and combined and harmonized ADRC registry and EHR data. For each data source and cognitive assessment, we report:
c. The number and proportion of patients experiencing first sustained cognitive worsening;
d. The average number of scores per patient;
e. The average duration between consecutive cognitive assessments in months.
f. CDR Global scores were only available from the ADRC registry.

Abbreviations: ADRC, Alzheimer's Disease Research Center; CDR, Clinical Dementia Rating; EHR, Electronic Health Record; MMSE, Mini-Mental State Examination; MoCA, Montreal Cognitive Assessment

**S-Table 8**

Predictive performance for patient-level classification of first sustained cognitive worsening in the gold-standard cohort. [a]

| Metric | CDR Global, mean [95% CI] | MMSE, mean [95% CI] | MoCA, mean [95% CI] |
|---|---|---|---|
| AUC | 0.831 [0.811 – 0.852] | 0.738 [0.718 – 0.758] | 0.770 [0.747 – 0.791] |
| Sensitivity | 0.750 [0.703 – 0.793] | 0.533 [0.496 – 0.568] | 0.620 [0.581 – 0.661] |
| PPV | 0.526 [0.480 – 0.573] | 0.701 [0.668 – 0.735] | 0.523 [0.484 – 0.563] |
| NPV | 0.916 [0.901 – 0.930] | 0.661 [0.640 – 0.684] | 0.857 [0.840 – 0.872] |

a. We derived discrimination thresholds at 80% specificity using 5-fold cross-validation with 1,000 bootstrap replicates in the gold-standard training set, then applied these thresholds in the test sets to classify patients as reaching (outcome=1) or not reaching (outcome=0) first sustained cognitive worsening. We report mean performance metrics with 95% confidence intervals.

Abbreviations: AUC, Area Under the Receiver Operating Characteristic Curve; CDR, Clinical Dementia Rating; MMSE, Mini-Mental State Examination; MoCA, Montreal Cognitive Assessment; NPV, Negative Predictive Value; PPV, Positive Predictive Value

**S-Figure 1**

Trends in average time-AUC for predicting first sustained cognitive worsening in gold-standard cohorts across cognitive assessments in the test set during each follow-up year.

Abbreviations: AUC, Area Under the Receiver Operating Characteristic Curve; CDR, Clinical Dementia Rating; MMSE, Mini-Mental State Examination; MoCA, Montreal Cognitive Assessment

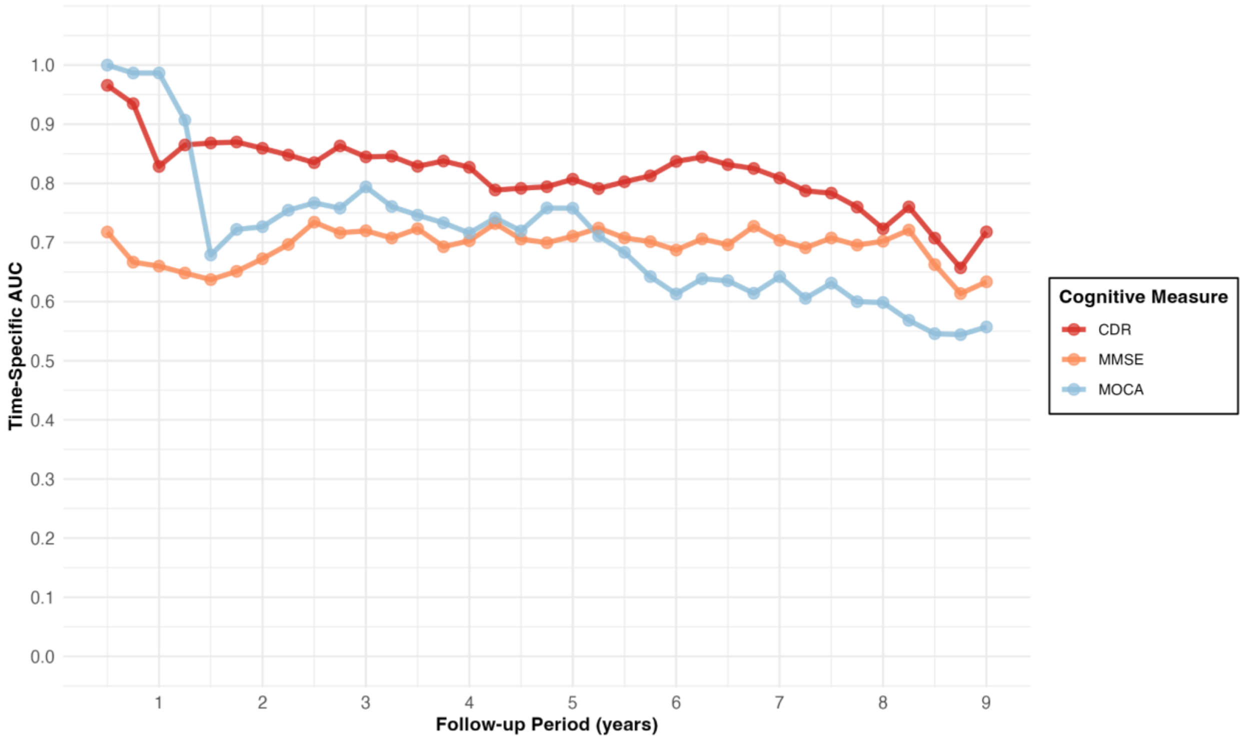

**S-Figure 2**

Trends in average time-AUC for predicting first sustained cognitive worsening in gold-standard cohorts across cognitive assessments in the test set, stratified by follow-up duration periods (<2 years, 2–5 years, or 5+ years). Mean time-specific AUC and error bars representing 95% confidence intervals are shown.

<u>Abbreviations</u>: AUC, Area Under the Receiver Operating Characteristic Curve; CDR, Clinical Dementia Rating; MMSE, Mini-Mental State Examination; MoCA, Montreal Cognitive Assessment

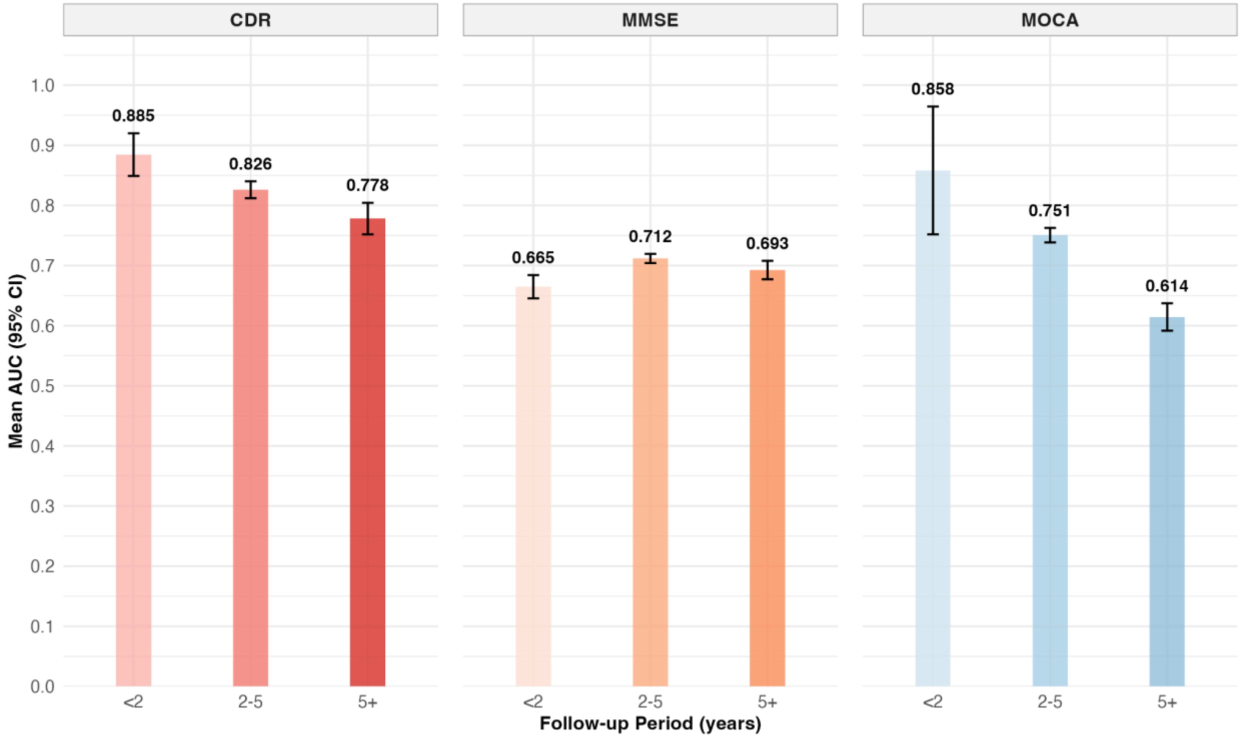

**S-Figure 3**

Performance of predicting first sustained cognitive worsening across cognitive assessments, stratified by baseline cognitive impairment severity. We report longitudinal model performance for CDR, MMSE, and MoCA scores. Baseline was defined as the first cognitive assessment score in the sustained cognitive worsening endpoint. We report performance for normal and mild baseline cognitive impairment severity, after excluding the moderate-to-severe group due to small sample size. Average time-Area Under the Receiver Operating Characteristic Curve (Average Time-AUC) quantifies discrimination as the mean of the time-specific AUC computed at each three-month interval during follow-up, with higher values indicating better model discrimination. Area Between Curves (ABC) quantifies calibration as the mean absolute difference between the observed and predicted cumulative incidence curves across all three-month intervals during follow-up, with lower values indicating better model calibration. Brier score measures overall predictive accuracy as the mean squared difference between the observed outcome and predicted probability of first sustained cognitive worsening, averaged across all three-month intervals during follow-up, with lower values indicating better model performance.

Abbreviations: AUC, Area Under the Receiver Operating Characteristic Curve; CDR, Clinical Dementia Rating; MMSE, Mini-Mental State Examination; MoCA, Montreal Cognitive Assessment

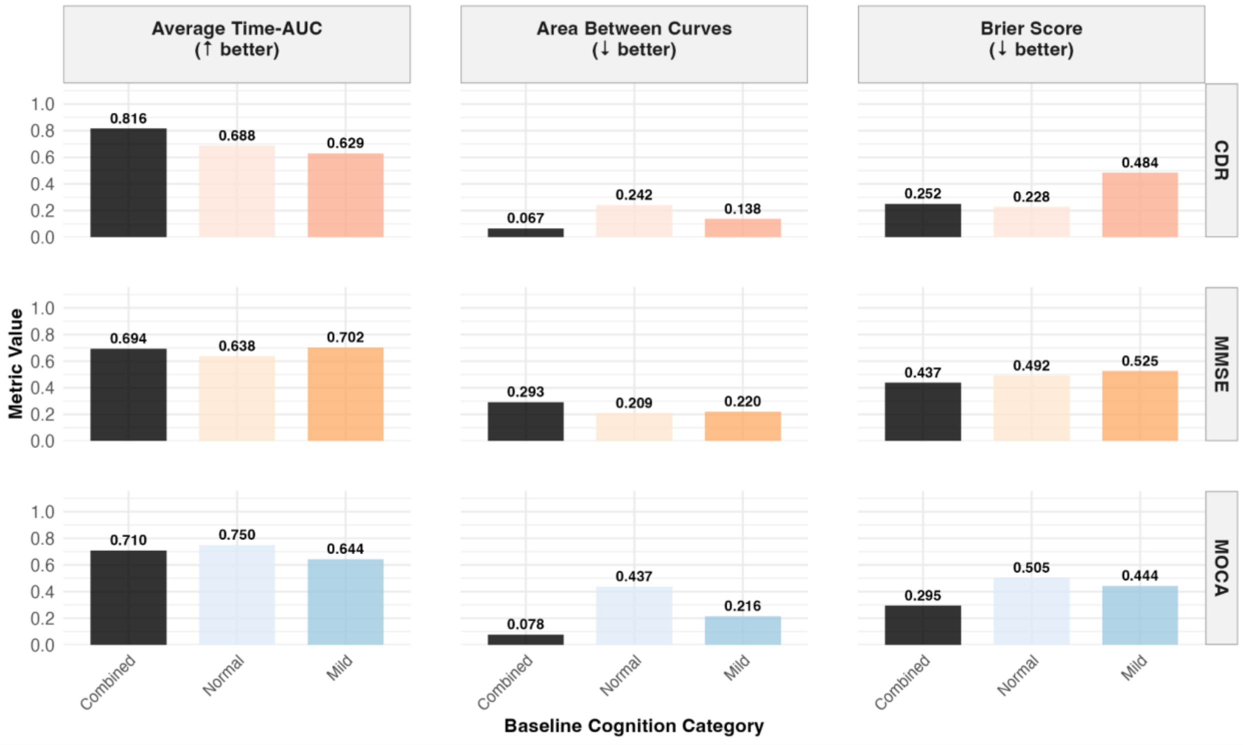

**S-Figure 4**

Performance of predicting first sustained cognitive worsening across cognitive assessments, stratified by cognitive assessment data source. We report longitudinal model performance for MMSE and MoCA. Data sources include ADRC registry data, EHR data, and combined and harmonized ADRC registry and EHR data. CDR Global scores were only available from the ADRC registry. Average time-Area Under the Receiver Operating Characteristic Curve (Average Time-AUC) quantifies discrimination as the mean of the time-specific AUC computed at each three-month interval during follow-up, with higher values indicating better model discrimination. Area Between Curves (ABC) quantifies calibration as the mean absolute difference between the observed and predicted cumulative incidence curves across all three-month intervals during follow-up, with lower values indicating better model calibration. Brier score measures overall predictive accuracy as the mean squared difference between the observed outcome and predicted probability of first sustained cognitive worsening, averaged across all three-month intervals during follow-up, with lower values indicating better model performance.

Abbreviations: ADRC, Alzheimer's Disease Research Center; AUC, Area Under the Receiver Operating Characteristic Curve; CDR, Clinical Dementia Rating; EHR, Electronic Health Record; MMSE, Mini-Mental State Examination; MoCA, Montreal Cognitive Assessment

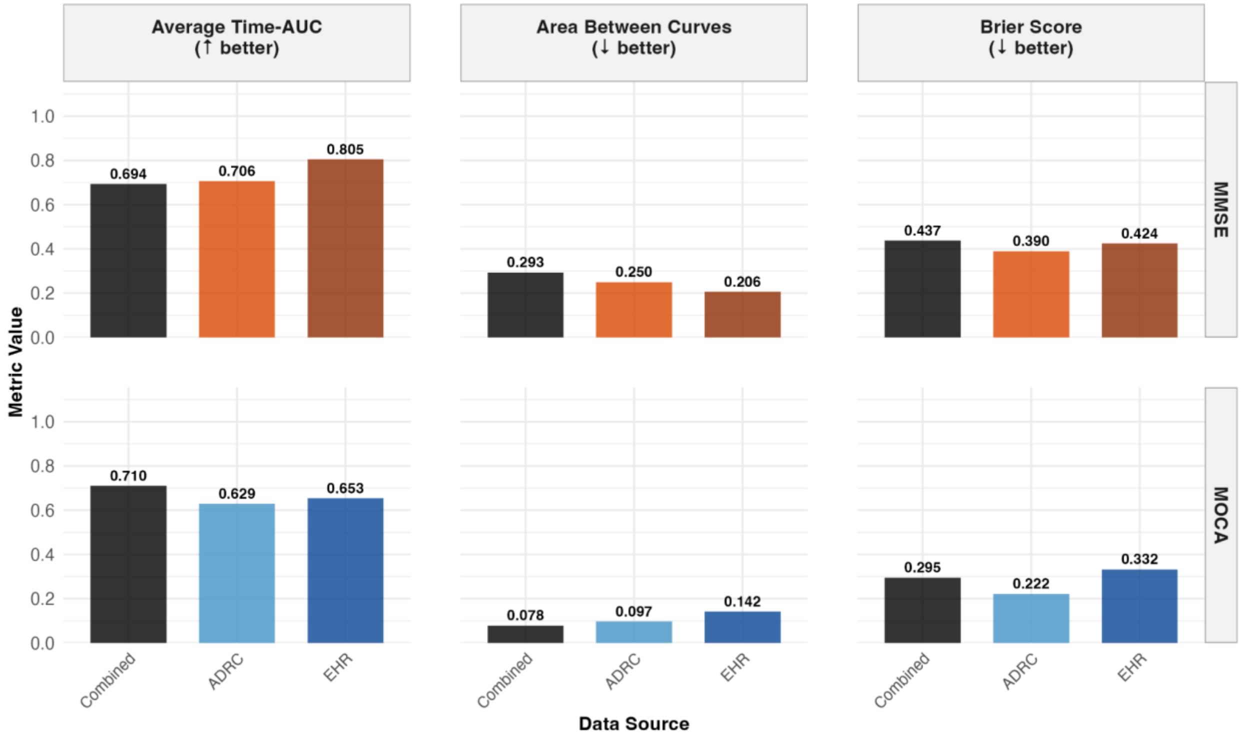

**S-Figure 5**

**Time-to-first sustained cognitive worsening stratified by *APOE*-ε4 carrier status in gold-standard cohorts.** We estimated time-to-first sustained cognitive worsening across all cognitive assessments using unadjusted, competing-risk Cox proportional hazards models, accounting for death as a competing event. Solid lines represent survival probability. Shaded regions represent 95% confidence intervals. The reference group for APOE-ε4 carrier status was non-carrier. **(A) CDR Global, gold-standard cohort** (n=628; 469 events). **(B) MMSE, gold-standard cohort** (n=619; 398 events). **(C) MoCA, gold-standard cohort** (n=376; 321 events).

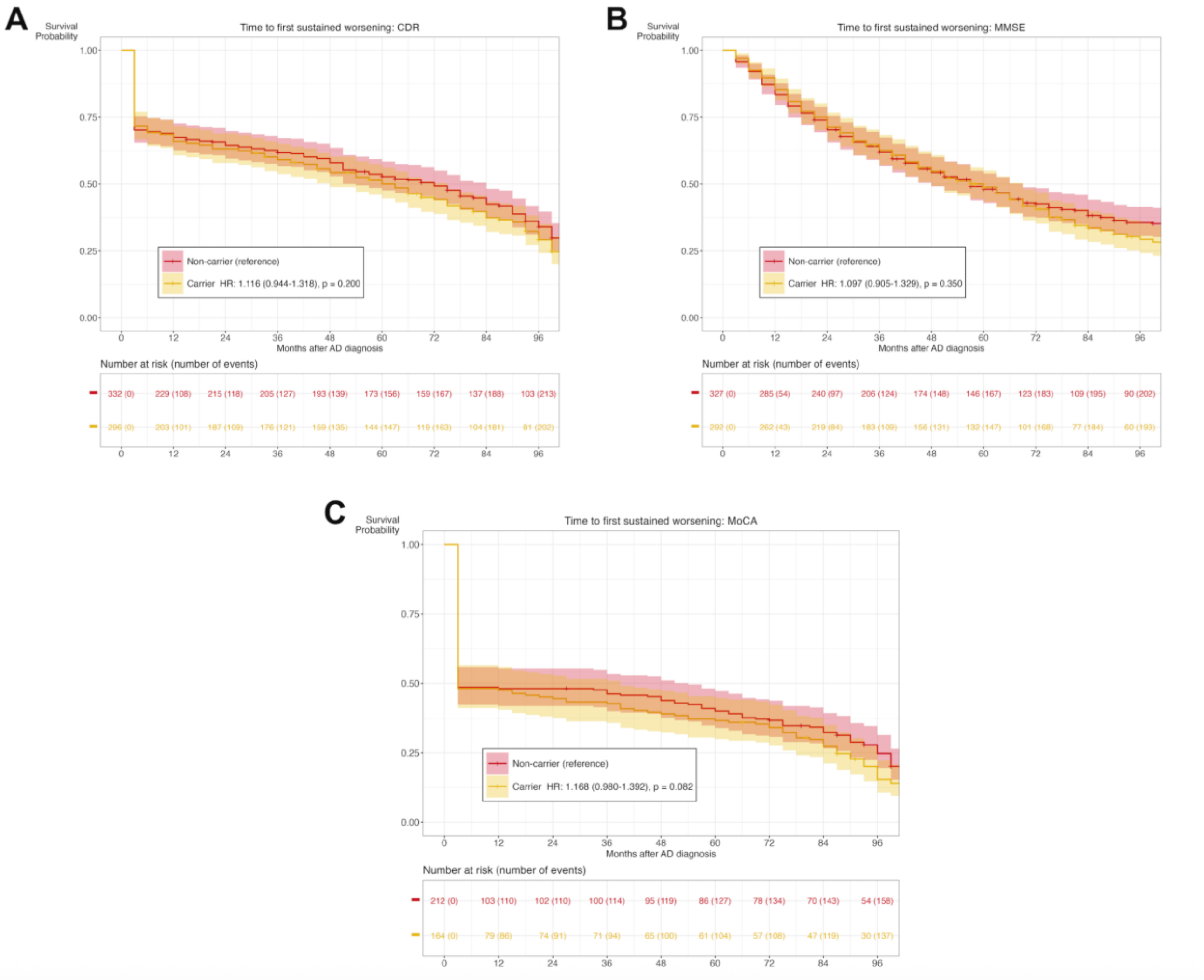

**S-Figure 6**

**Time-to-first sustained cognitive worsening stratified by *APOE*-ε4 carrier status in imputation cohorts.** We estimated time-to-first sustained cognitive worsening across all cognitive assessments using unadjusted, competing-risk Cox proportional hazards models, accounting for death as a competing event. Solid lines represent survival probability. Shaded regions represent 95% confidence intervals. The reference group for APOE-ε4 carrier status was non-carrier. **(A) CDR Global, imputation cohort** (n=400; 210 events). **(B) MMSE, imputation cohort** (n=409; 366 events). **(C) MoCA, imputation cohort** (n=652; 274 events).

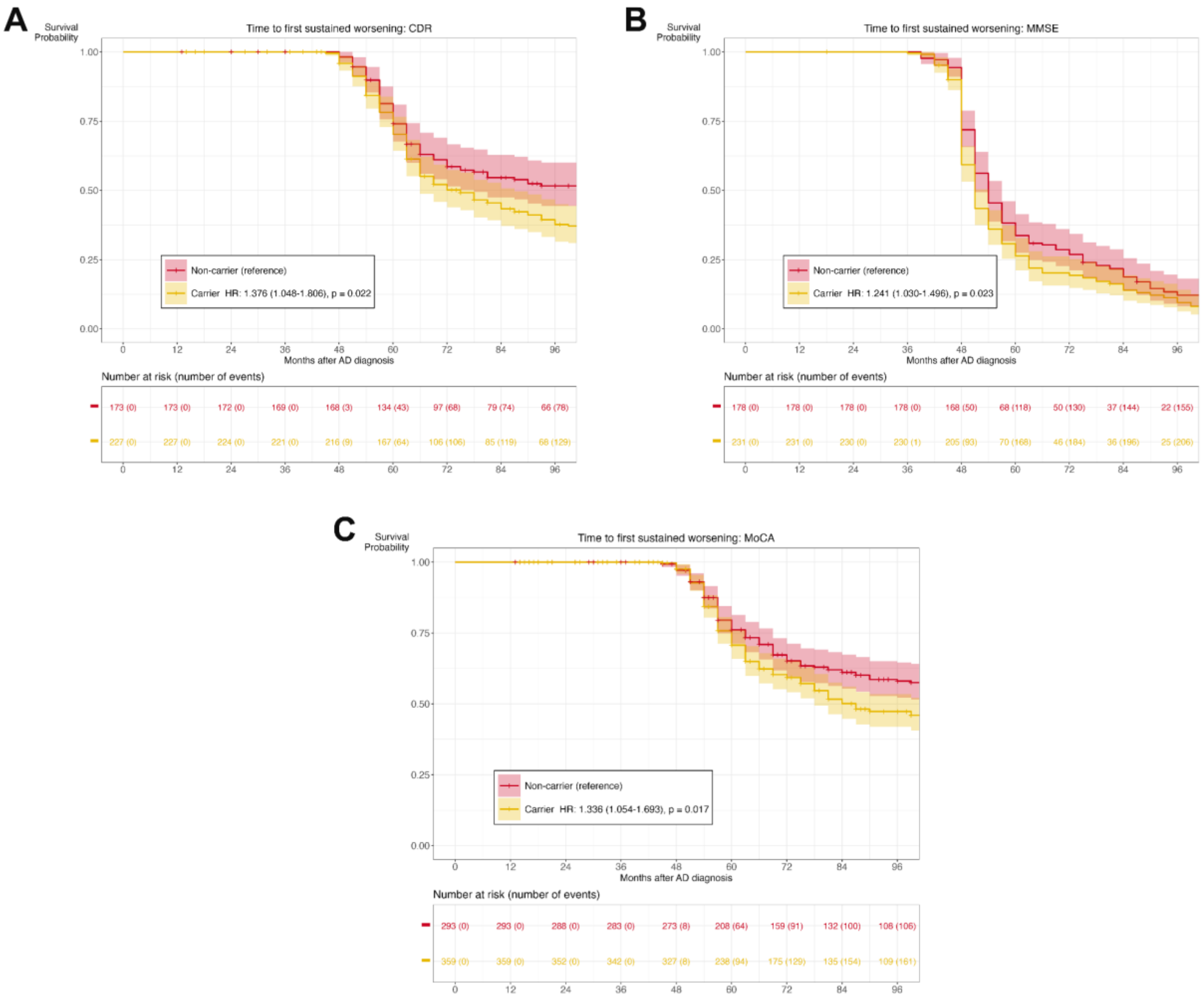

**S-Figure 7**

**Selection of optimal number of patient clusters using the silhouette method.** We performed k-means clustering of knowledge-graph-weighted baseline EHR feature embeddings and selected the optimal number of clusters (k) using the silhouette method. The silhouette width quantifies, for each patient, how well they fit their assigned cluster compared to the nearest neighboring cluster, ranging from −1 (closer to a neighboring cluster than to the assigned one) through 0 (on the boundary) to +1 (well-matched to the assigned cluster). We selected the *k* that maximized the mean silhouette width across all patients.

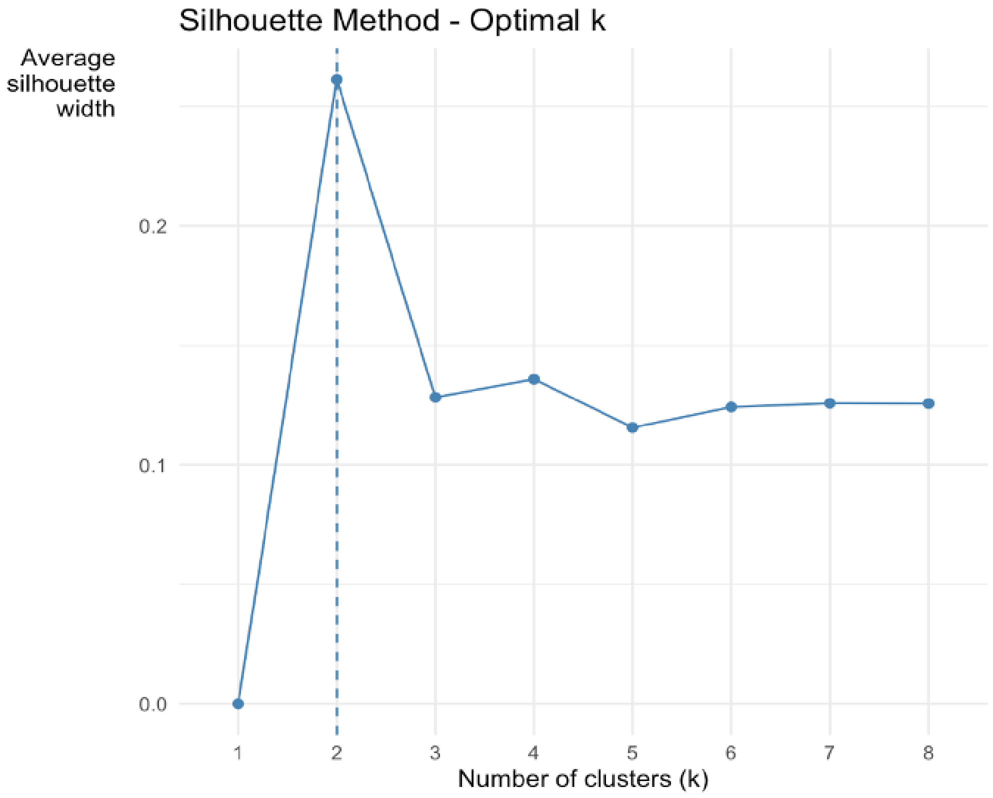

**S-Figure 8**

**Time-to-first sustained cognitive worsening stratified by baseline patient clustering in gold-standard cohorts.** We excluded patients with first cognitive worsening or <24 months of EHR data prior to AD diagnosis. We clustered the remaining patients into two subgroups using k-means clustering (k=2) of knowledge-graph-weighted EHR feature embeddings, adjusted for healthcare utilization. We estimated time-to-first sustained cognitive worsening using unadjusted, competing-risk Cox proportional hazards models accounting for death as a competing event. Solid lines represent survival probability. Shaded regions represent 95% confidence intervals. Reference subgroup: Cluster 1. **(A) CDR Global, gold-standard cohort** (n=155; 136 events). **(B) MMSE, gold-standard cohort** (n=192; 159 events). **(C) MoCA, gold-standard cohort** (n=301; 272 events).

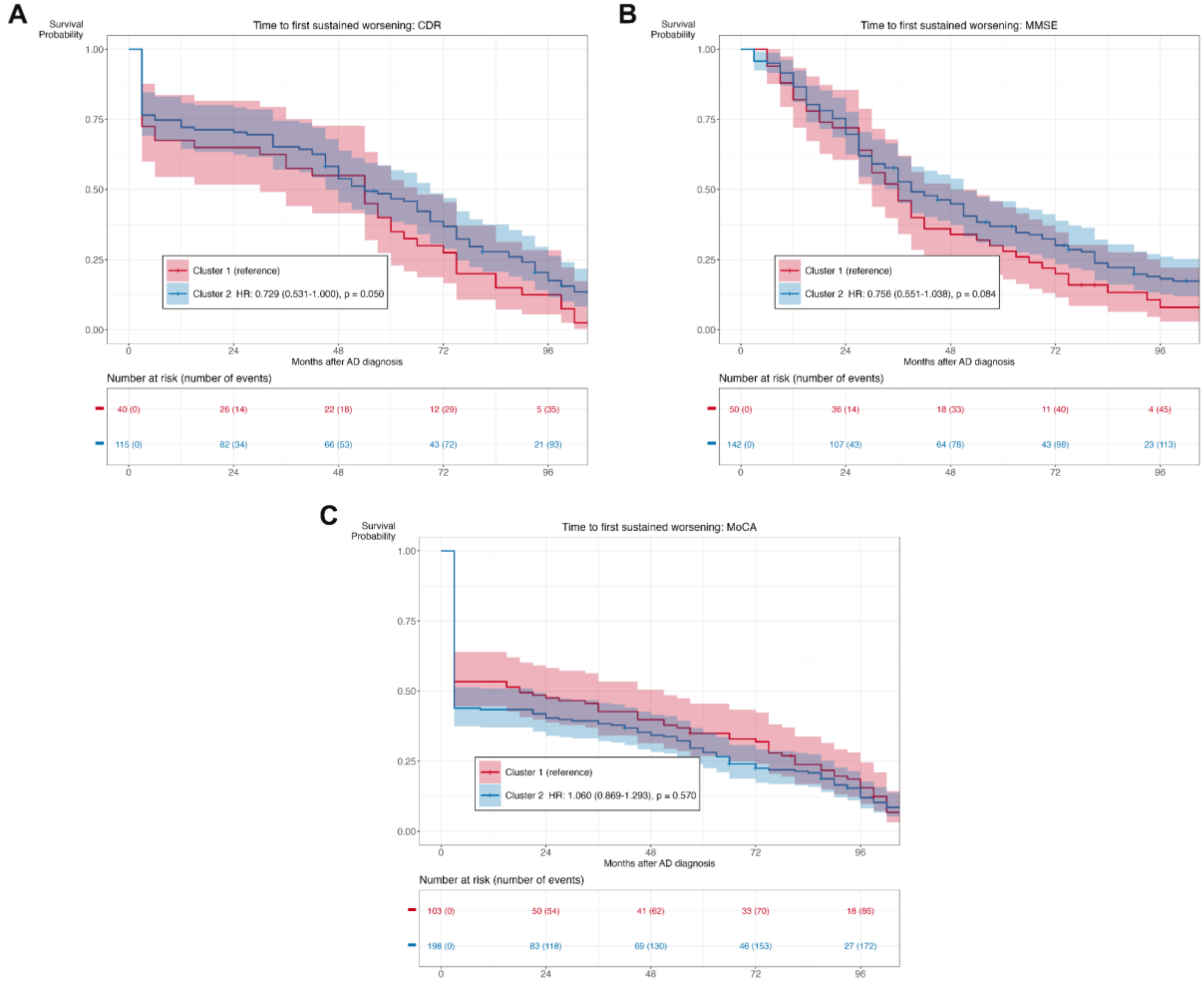

**S-Figure 9**

**Time-to-first sustained cognitive worsening stratified by baseline patient clustering in imputation cohorts.** We excluded patients with first cognitive worsening or <24 months of EHR data prior to AD diagnosis. We clustered the remaining patients into two subgroups using k-means clustering (k=2) of knowledge-graph-weighted EHR feature embeddings, adjusted for healthcare utilization. We estimated time-to-first sustained cognitive worsening using unadjusted, competing-risk Cox proportional hazards models accounting for death as a competing event. Solid lines represent survival probability. Shaded regions represent 95% confidence intervals. Reference subgroup: Cluster 1. **(A) CDR Global, imputation cohort** (n=14,092; 6,268 events). **(B) MMSE, imputation cohort** (n=14,055; 12,527 events). **(C) MoCA, imputation cohort** (n=13,946; 6,054 events).

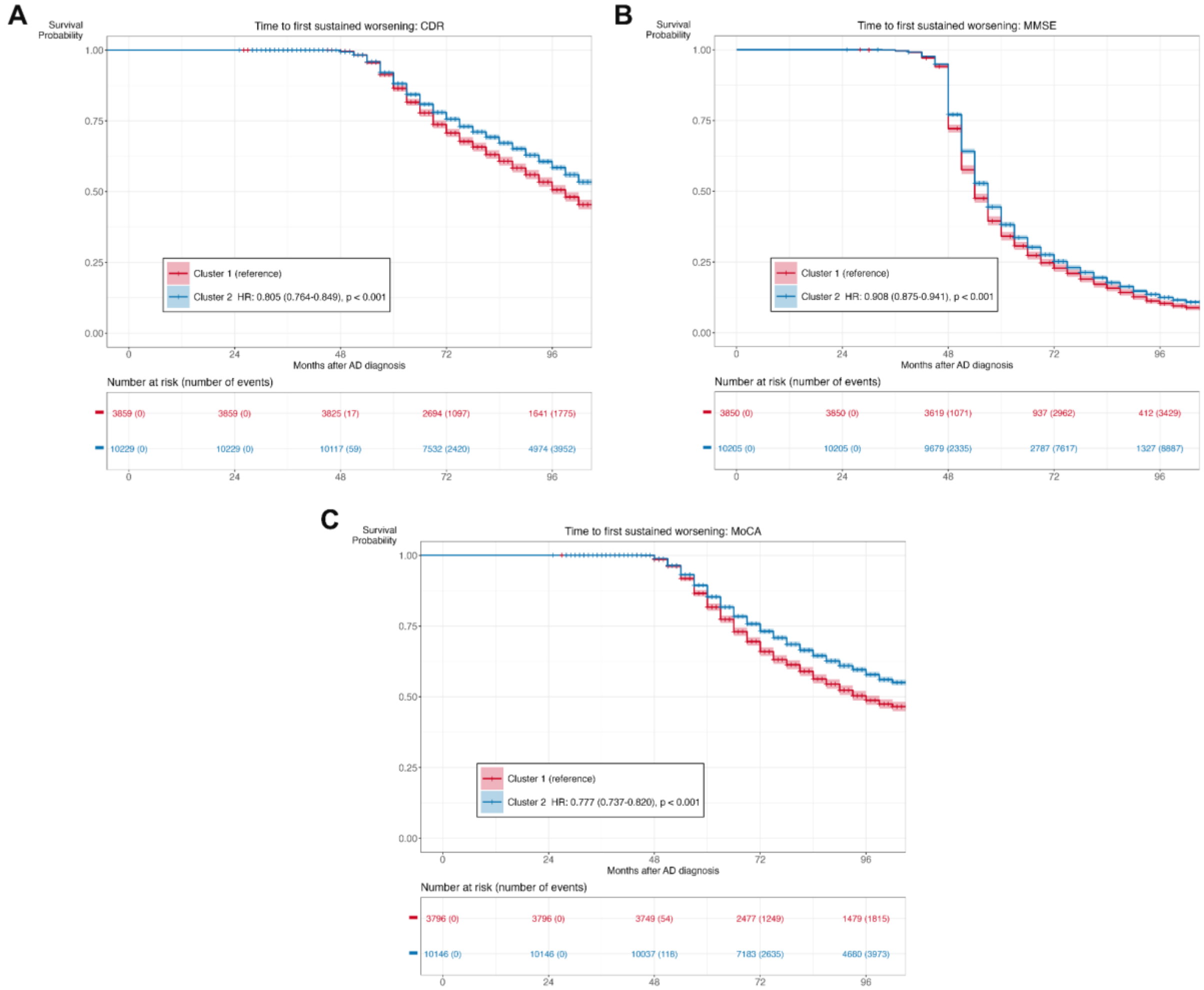

**S-Figure 10**

**Features distinguishing patient subgroups derived from k-means clustering at AD diagnosis.** The word cloud illustrates the features with the highest importance distinguishing Cluster 1 (red) from Cluster 2 (blue). Text size denotes the relative importance of each feature.

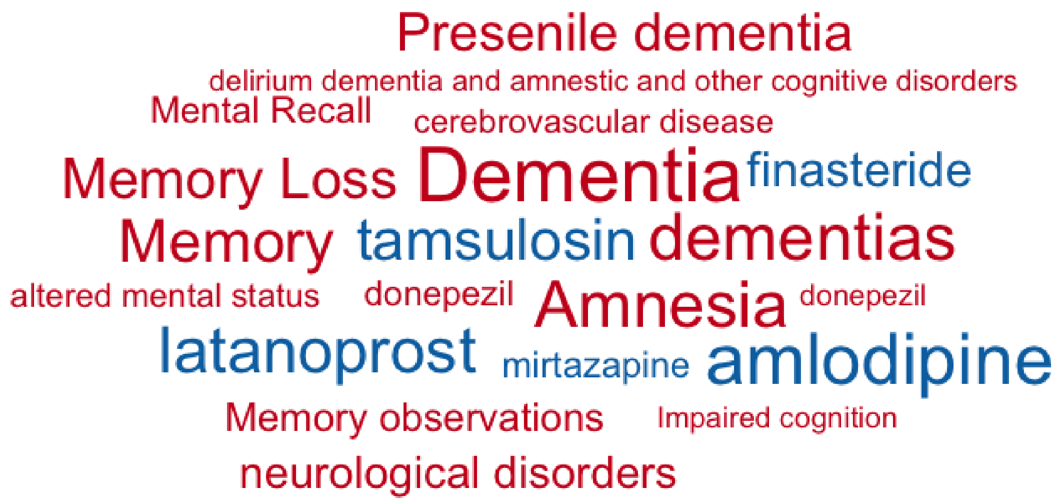
